# A data-driven dietary pattern anchored to slower epigenetic aging is associated with a spectrum of aging-related health outcomes

**DOI:** 10.64898/2026.02.23.26346925

**Authors:** Sirui Lai, Li Zhang, Jie Yu, Min Wu, Guoping Peng, Geng Zong, Hao Ma, Changzheng Yuan, Hui Chen, Benyan Luo

## Abstract

Diet is an essential factor influencing biological aging, yet few exsiting dietary indices were specifically developed to target biological aging. We developed a data-driven food-based Empirical Dietary Index for Slower Epigenetic Aging (EDISEA) in the US Health and Retirement Study (HRS, n=7,398), which predicted deceleration of GrimAge, an established DNA methylation-based epigenetic clock. Participants in the highest versus lowest EDISEA quintile had 4.65-year deceleration in GrimAge (P value <0.001). We externally validated EDISEA in an independent US cohort (n=23,830), where it showed consistent associations with several epigenetic clocks and lower all-cause mortality risk. In HRS and a UK aging cohort (n=4,895), EDISEA was associated with lower risks of several aging-related diseases and functional limitations. Outcome-wide analyses in the UK Biobank (n=187,035), together with integrative proteomic, metabolic, and neuroimaging assessments, revealed biological signatures of EDISEA implicating broad vascular, inflammatory, metabolic, and brain-structural pathways through which EDISEA was associated with biological aging. EDISEA provides a scalable, biologically anchored tool to inform the development of precision nutrition strategies aimed at slowing epigenetic aging and mitigating aging-related disease burden.

## Introduction

Aging is a complex, multifactorial process marked by the progressive decline of physiological integrity and increasing vulnerability to chronic diseases^1,2^. Substantial inter-individual variation in the pace of biological aging has motivated efforts to identify modifiable determinants that may delay aging-related deterioration^3–5^. Among emerging biomarkers, epigenetic clocks derived from DNA methylation profiles have shown robust associations with morbidity, mortality, and aging-related functional impairments, and are increasingly recognized as integrative measures of biological aging^6–9^.

Importantly, epigenetic aging appears modifiable as a promising intermediate target for achieving healthy aging^10,11^. Diet has broad relevance to biological aging^12–14^, and a large body of observational and interventional research has linked dietary factors to longevity, cardiometabolic health, and neurodegenerative outcomes^15–18^. As of now, most prior studies have focused on individual nutrients or a priori-defined dietary scores, such as the Mediterranean, DASH, or Mediterranean-DASH Intervention for Neurodegenerative Delay (MIND) diets. For example, in the Framingham Heart Study Offspring cohort, higher DASH diet quality was associated with lower epigenetic age acceleration across multiple DNAm-based clocks, which partially mediated the association between DASH and all-cause dementia and mortality^19,20^. Similarly, in the Women’s Health Initiative, higher overall diet quality was consistently associated with younger epigenetic aging profiles^21^. However, three gaps remain. First, these hypothesis-driven approaches may not fully capture the complexity and synergistic nature of dietary intake in epigenetic aging. Recent advances in high-dimensional data analysis enabled the derivation of data-driven and pathway-informed dietary patterns that more comprehensively reflect the biological processes modulated by diet. Empirically derived dietary indices, such as the empirical dietary index for hyperinsulinemia (EDIH) and inflammation (EDIP), have been constructed based on their associations with intermediate metabolic and inflammatory phenotypes and have demonstrated relevance across multiple health outcomes^22–26^. Such methodological advances have great potential to be integrated into studies of dietary influences on epigenetic aging and enable systematically examination the biological pathways that may underlie these associations. Second, existing analyses have typically focused on narrow sets of outcomes, and the broader health relevance and consistency of diet-epigenetic aging associations across aging-related phenotypes remain unclear. Third, prior work rarely examined biological pathways that may connect whole-diet patterns to epigenetic aging. Addressing these gaps is essential for an empirically grounded understanding of how habitual diet may influence the pace of epigenetic aging and inform dietary intervention strategies to achieve healthy aging.

In this study, we developed a data-driven dietary pattern explicitly anchored to epigenetic aging, named Empirical Dietary Index for Slower Epigenetic Aging (EDISEA), with the aim of identifying a dietary pattern associated with slower epigenetic aging. We validated this pattern in an independent sample and systematically evaluated its associations with a broad spectrum of aging-related outcomes using an outcome-wide analytical framework^27^. To enhance biological interpretability, we further explored potential mechanistic pathways linking the identified dietary pattern to epigenetic aging (**Figure 1**). This integrative approach aims to understand how habitual diet relates to epigenetic aging and to provide an empirically grounded dietary framework with potential relevance for strategies to slow epigenetic aging.

**Figure 1.**
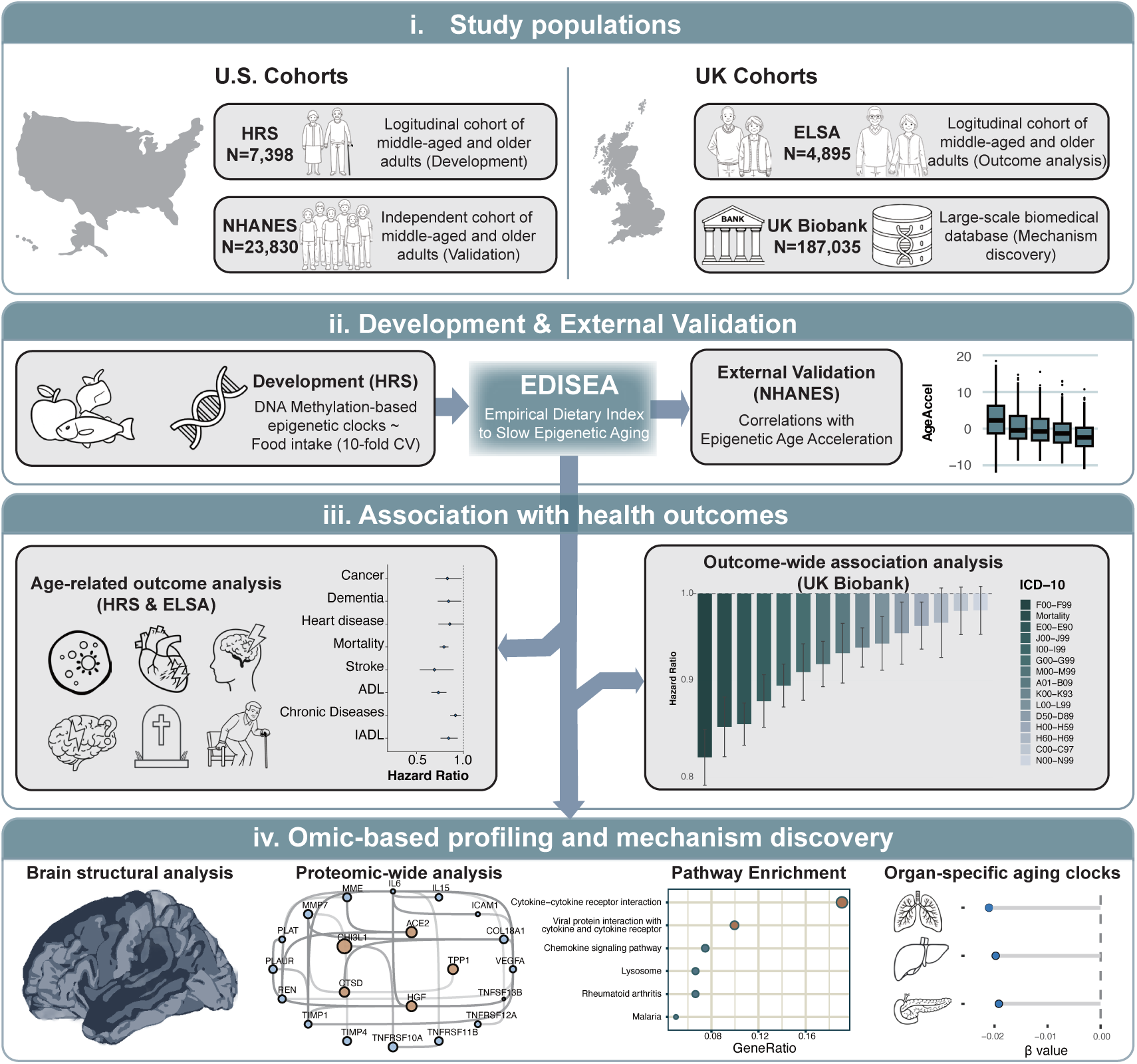
Design of the study. **Footnote:** (i) Study populations. Analyses were conducted in two U.S. cohorts (HRS, N=7,398; NHANES, N=23,830) and two U.K. cohorts (ELSA, N=4,895; UK Biobank, N=187,035). HRS was used for index development, NHANES for external validation, HRS and ELSA for primary aging-related outcome analyses, and the UK Biobank for outcome-wide association screening and mechanistic profiling. (ii) Development and external validation. An Empirical Dietary Index for Slower Epigenetic Aging (EDISEA) was developed in HRS by relating harmonized food-group intakes to DNAm-based epigenetic aging measures using cross-validated modeling, and then externally validated in NHANES by testing associations of EDISEA with epigenetic age acceleration. (iii) Association with health outcomes. Associations between EDISEA and major aging-related outcomes (including mortality, cancer, dementia, cardiovascular diseases, stroke, and functional impairment) were evaluated in prospective cohorts, and an outcome-wide association analysis across ICD-10-defined endpoints was performed in the UK Biobank. (iv) Omic-based profiling and mechanism discovery. Mechanistic analyses leveraged brain structural imaging, plasma proteome-wide association testing, pathway enrichment, protein-protein interaction networks, and organ-specific aging clocks to contextualize biological pathways linking EDISEA to aging-related outcomes.

## Results

### Food-group intake and epigenetic aging

Baseline characteristics of the analytic samples were summarized in **Table S1**. The current analyses included 7,398 participants from the US Health and Retirement Study (HRS, mean age of 68.7 years, SD 11.0; 41% male) as the development cohort, 23,830 from US NHANES as the external validation cohort (mean age of 62.7 years, SD 11.2; 48% male). Two UK-based cohorts were further included to explore the health outcome implications, including 4,895 from the English Longitudinal Study of Ageing (ELSA, mean age of 68.0 years, SD 7.9; 45% male), and 187,035 from the UK Biobank (mean age of 56.1 years, SD 8.0; 45% male). The cohorts had substantial diversity in ethnicity, educational attainment and income distributions.

In the HRS, we first explored the associations of 38 food groups, amalgamated according to a widely accepted framework^28^, with GrimAge acceleration (GrimAgeAccel), a well-established epigenetic age clock based on DNA methylation that shows the strongest associations with mortality and aging-related health outcomes^7^. In the fully adjusted linear regression models, eight food groups showed significant associations with GrimAgeAccel after false disvcovery rate correction (P value <0.05 after FDR correction), including four related to faster GrimAgeAccel, and four to slower GrimAgeAccel (**Figure 2a)**. The standardized GrimAgeAccel differences comparing 90^th^ to 10^th^ percentile of intake were 0.115 (95% CI 0.032 - 0.198) for processed meat, 0.079 (95% CI 0.047 - 0.112) for beer, 0.058 (95% CI 0.002 - 0.113) for artificial sweetened beverages, and 0.037 (95% CI 0.019 - 0.054) for liquor. Conversely, tea (β = -0.078, 95% CI -0.141 - -0.014), eggs (β = -0.089, 95% CI -0.162 - -0.015), fruits (β = -0.098, 95% CI -0.186 - -0.010), and whole grains (β = -0.103, 95% CI -0.187 - - 0.018) were associated with slower GrimAgeAccel. In the restricted cubic spline analyses, no food group showed evidence of non-linear associations (P value for non-linearity >0.05 after FDR correction). Therefore, linear specifications were applied in subsequent analyses.

**Figure 2.**
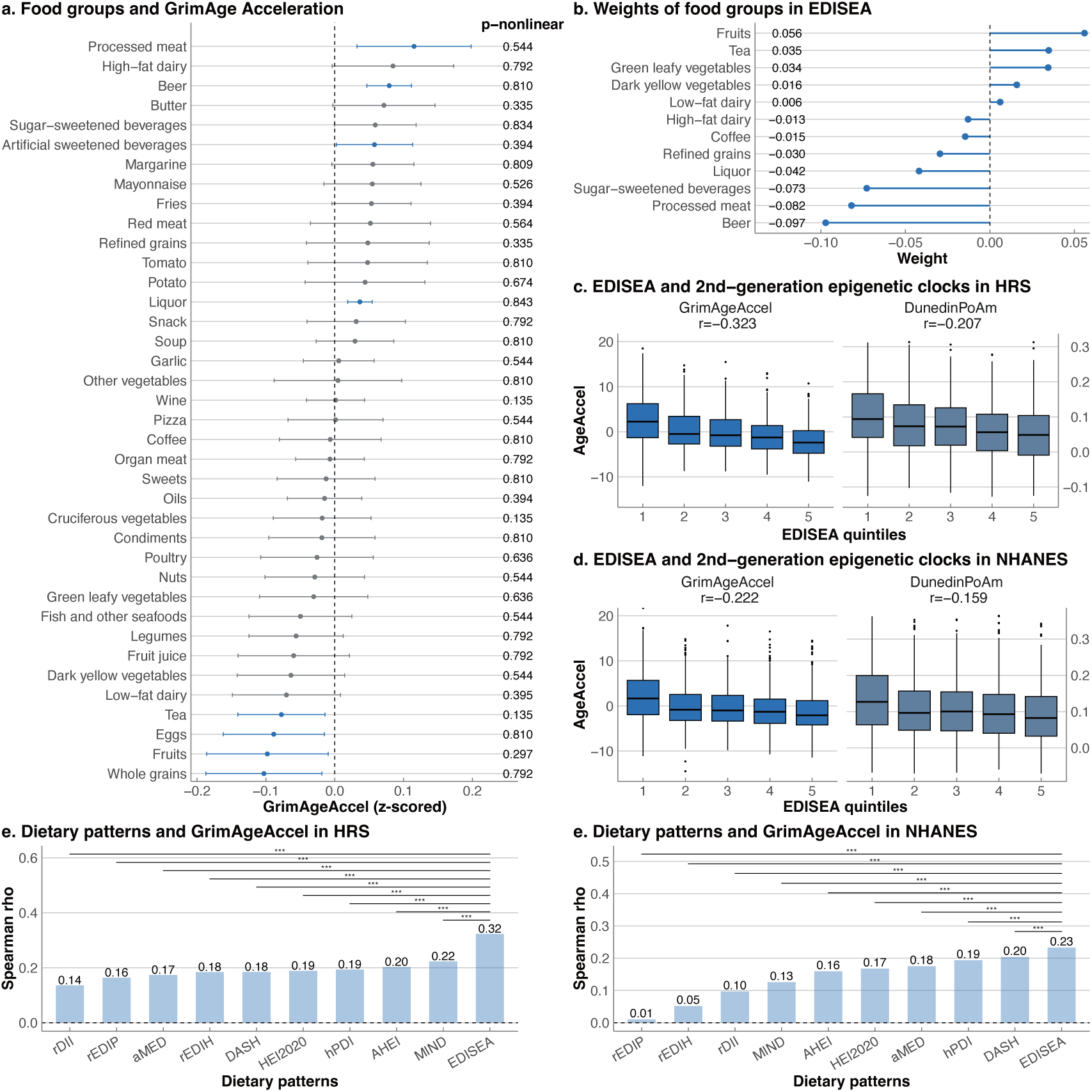
Food-wide associations with GrimAge and development and validation of the EDISEA. **Footnote: a**, Food groups and GrimAge acceleration in HRS. Associations of individual food-group intakes with standardized GrimAgeAccel were estimated using multivariable linear regression. Effect estimates are shown as regression coefficients (β) and 95% confidence intervals per 90^th^ to 10th percentile difference in intake for each food group. Models were adjusted for age, sex, ethnicity, education qualification, income, smoking status, alcohol consumption status, physical activity, BMI, and total energy intake. Points are colored by nominal P value (<0.05 vs ≥0.05). The vertical dashed line indicates the null (β=0). **b**, Weights of food groups in EDISEA. Food-group weights were derived from a cross-validated elastic-net regression model in HRS, fit to standardized food-group intakes. Only food groups with non-zero weights are shown. The sign convention was oriented such that higher EDISEA reflects a dietary pattern associated with slower epigenetic aging in the derivation cohort. **c**, EDISEA and second-generation epigenetic aging measures in HRS. Distributions of epigenetic aging measures across EDISEA quintiles are shown as boxplots; boxplots show median (center line), interquartile range (box), and 1.5×IQR whiskers; points denote individual participants. Spearman correlations (ρ) between continuous EDISEA and each aging metric are indicated. For GrimAgeAccel, the y axis denotes age acceleration in years. For DunedinPoAm, values are displayed on the right y axis in the original DunedinPoAm scale (deviation from one biological year per chronological year, for example, 0.10 indicates 10% faster and −0.10 indicates 10% slower biological aging). **d**, External validation in NHANES. The same EDISEA scoring approach was applied to harmonized NHANES dietary data, and associations with epigenetic aging measures were evaluated analogously; boxplots show epigenetic aging measures across EDISEA quintiles with Spearman ρ values. Age acceleration measures are the same as panel **c**. **e,f,** Bar plots show Spearman rank correlations between each dietary pattern score and GrimAgeAccel in HRS (e) and NHANES (f). DII, EDIH and EDIP were reversed to rDII, rEDIH, rEDIP respectively so that higher values always reflect healthier dietary patterns. Statistical significance for pairwise comparisons of correlation magnitudes verses EDISEA was assessed using nonparametric bootstrap resampling. ***p<0.001.

### Development and external validation of EDISEA

We used 10-fold cross-validated elastic-net regression model to predict GrimAgeAccel using standardized food-group intake levels **(Figure 2b, and Table S2)**. The elastic-net regression model identified twelve food groups with non-zero weights, which were used to construct the Empirical Dietary Index for Slower Epigenetic Aging (EDISEA) and reflect joint contributions of dietary factors to epigenetic aging. Foods with positive weights contributing to higher EDISEA included fruits (0.056), tea (0.035), green leafy vegetables (0.034), dark yellow vegetables (0.016), and low-fat dairy (0.006). Foods with negative weights contributing to lower EDISEA included beer (−0.097), processed meat (−0.082), sugar-sweetened beverages (−0.073), liquor (−0.042), refined grains (−0.030), coffee (−0.015), and high-fat dairy (−0.013). These weights aligned with the food-wide association results as described above, and the selected food groups were stable across cross-validation folds and penalty parameters. The same approach was also applied to predict DunedinPoAm as sensitivity analyses **(Table S3)**. EDISEA showed moderate correlations with several established dietary indices, such as alternate Mediterranean diet (Spearman’s ρ = 0.55 in HRS and 0.42 in NHANES), the brain-health-targeting MIND diet (ρ = 0.57 in HRS and 0.32 in NHANES), and the data-driven reversed empirical dietary inflammatory pattern (rEDIP) (ρ = 0.35 in HRS and 0.10 in NHANES), reflecting similar but distinct constructs (**Figure S2**).

In internal validation within HRS, higher EDISEA was associated with more favorable profiles across two second-generation epigenetic aging metrics **(Figure 2c, and Table S4)**, showing graded differences across EDISEA quintiles and inverse Spearman correlations with GrimAgeAccel (ρ = −0.323) and DunedinPoAm (ρ = −0.207). Notably, the mean GrimAgeAccel decreased monotonically from 2.62 years in the lowest EDISEA quintile to −2.03 years in the highest quintile, corresponding to an approximately 4.6-year difference between the extreme quintiles. Applying the same EDISEA scoring approach to harmonized NHANES dietary data yielded consistent external validation **(Figure 2d, and Table S4)**, again showing inverse correlations with GrimAgeAccel (ρ = −0.222) and DunedinPoAm (ρ = −0.159). EDISEA was also positively correlated with DNAm-estimated telomere length (ρ = 0.100), consistent with a more favorable epigenetic aging profile, supporting the generalizability of EDISEA across cohorts with distinct dietary assessment contexts. In NHANES, GrimAgeAccel also decreased monotonically across EDISEA quintiles (mean: 2.20 years in Q1, −1.17 years in Q5), corresponding to an approximately 3.4-year deceleration. Notably, the EDISEA showed stronger correlations with the epigenetic age acceleration measures compared with other established dietary indices in both the development and validation cohorts (**Figure 2e, f**).

### EDISEA and aging-related health outcomes

After adjustment for sociodemographic, lifestyle, and clinical factors, higher EDISEA was related to lower risk of mortality **(Figure 3a, b, and Table S5)**, with a 90^th^ to 10^th^ percentile difference in EDISEA associated with 22% lower mortality risk in HRS (hazard ratio [HR]: 0.78, 95% CI 0.69 - 0.89; 1,642 deaths over 69,379 person-years) and 21% in NHANES (HR: 0.79, 95% CI 0.74 - 0.84; 5,982 deaths over 234,439 person-years). The pooled HR was 0.79 (95% CI 0.74 - 0.83) **(Figure 3c)**. Associations were generally similar across study subgroups defined by age, gender, education attainment, income, smoking status, and BMI, with evidence of stronger associations among population aged below 65 years (P value for interaction <0.05) **(Table S6)**. Findings remained similar in sensitivity analyses by excluding deaths within the first two years of follow-up and excluding participants with baseline functional limitations **(Table S7).**

**Figure 3.**
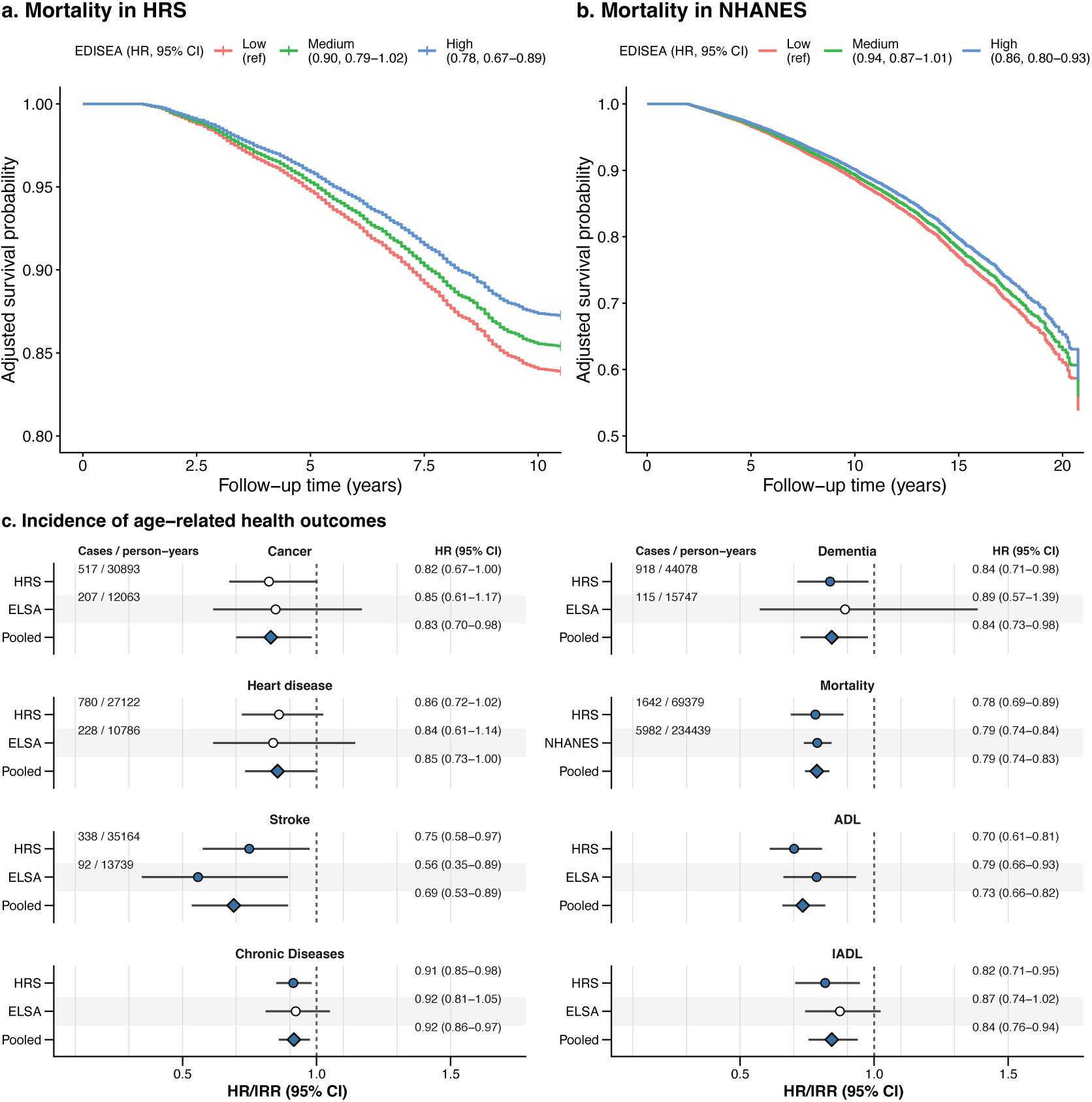
Associations of EDISEA with mortality and other aging-related health outcomes. **Footnote: a**,**b**, Mortality. Adjusted survival curves for mortality according to tertiles of EDISEA (low, medium, high) in HRS (**a**) and NHANES (**b**). Curves were generated from multivariable Cox proportional hazards models by predicting survival at the mean (continuous covariates) or reference category (categorical covariates) of covariates. Follow-up time was calculated from baseline assessment to date of death or censoring. **c**, Incidence of aging-related health outcomes. Forest plots show hazard ratios (HRs) or incidence rate ratios (IRRs) comparing EDISEA (scaled to the 90^th^ to 10^th^ percentile difference within cohort) with incident outcomes, with cohort-specific estimates for HRS, ELSA, and (where available) NHANES, and pooled estimates from random-effects meta-analysis. Points indicate effect estimates and horizontal lines 95% confidence intervals; diamonds denote pooled estimates. Numbers at left indicate cases and person-years for each cohort and outcome. The vertical dashed line denotes the null (HR/IRR=1). Time-to-event outcomes (cancer, dementia, heart disease, stroke, mortality) were analyzed using Cox proportional hazards regression; count-type outcomes (ADL, IADL, and total chronic disease incidence, as applicable) were analyzed using log-linked Poisson regression (with an offset for follow-up time when modeling incidence rates). All models were adjusted for a harmonized set of baseline covariables including age, sex, ethnicity, education qualification, income, smoking status, alcohol consumption status, physical activity, BMI, and total energy intake. ADL, Activities of Daily Living; IADL, Instrumental Activities of Daily Living.

In addition, higher EDISEA was associated with lower incidence of a spectrum of aging-related outcomes in prospective analyses of HRS and ELSA **(Figure 3c)**, including cancer (pooled HR: 0.83, 95% CI 0.70 - 0.98), dementia (pooled HR: 0.84, 95% CI 0.73 - 0.98), heart disease (pooled HR: 0.85, 95% CI 0.73 - 1.00), and stroke (pooled HR: 0.69, 95% CI 0.53 - 0.89). Consistently, higher EDISEA was also associated with lower rates of functional limitation and multimorbidity burden, with pooled incidence rate ratios being 0.73 (95% CI 0.66 - 0.82) for ADL limitation, 0.84 (95% CI 0.76 - 0.94) for IADL limitation, and 0.92 (95% CI 0.86 - 0.97) for total chronic disease incidence **(Figure 3c)**. Higher EDISEA was also associated with slower Pace of Aging (PoA) measure based on 9 biomarkers in HRS with no evidence of non-linearity (P-overall = 0.043, P-nonlinear = 0.239) **(Figure S3)**.

In the UK Biobank, higher EDISEA was associated with lower mortality risk (HR = 0.85, 95% CI 0.82 - 0.88) and multiple incident ICD-10-defined outcomes, with 42 out of 59 associations showing statistical significance after FDR correction **(Figure 4, and Table S5)**. The clearest signals clustered in major aging-related health outcomes, including cardiometabolic and vascular outcomes (e.g., diabetes: HR = 0.76, 95% CI 0.72 - 0.80; hypertension: HR = 0.84, 95% CI 0.82 - 0.87; ischaeomic heart disease: HR = 0.87, 95% CI 0.84 - 0.91; stroke: HR = 0.85, 95% CI 0.80 - 0.92; heart failure: HR = 0.86, 95% CI 0.81 - 0.91) and cardiopulmonary disease (chronic obstructive pulmonary disease: HR = 0.80, 95% CI 0.76 - 0.85). In addition, several aging-linked cancers and end-organ outcomes showed consistent inverse associations (e.g., lung cancer: HR = 0.79, 95% CI 0.71 - 0.89; oesophagus cancer: HR = 0.69, 95% CI 0.57 - 0.85; colorectal cancer: HR = 0.86, 95% CI 0.78 - 0.94; liver diseases: HR = 0.72, 95% CI 0.68 - 0.76; renal failure: HR = 0.81, 95% CI 0.78 - 0.84). By contrast, EDISEA showed no significant association (P value >0.05) with external-cause mortality (ICD-10: V01-V97, W00-W99, X00-X49, X58-X99, Y00-Y34, Y40-Y69, Y83-Y89), which was specified as the negative control outcome. Therefore, the associations of EDISEA with aging-related health outcomes were less likely to be systematically driven by outcomes unrelated to biological aging, suggesting limited influence from residual socioeconomic confounding.

**Figure 4.**
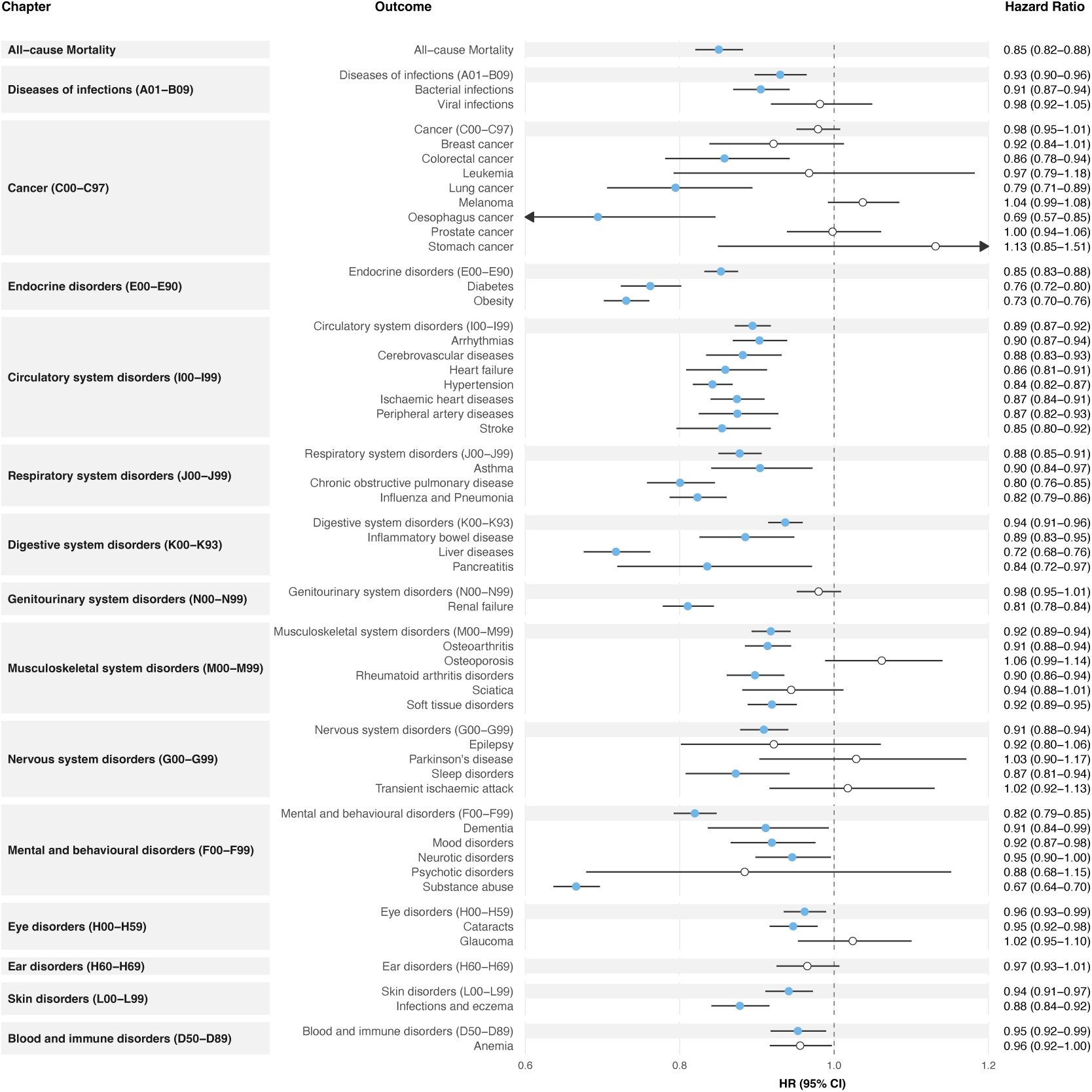
Outcome-wide association analysis of the EDISEA in the UK Biobank. **Footnote**: Forest plot showing hazard ratios (HRs) and 95% confidence intervals for the association between EDISEA and incident ICD-10-defined health outcomes in UK Biobank. Effect estimates were obtained from Cox proportional hazards models with follow-up time as the underlying time scale, with baseline defined as the date of the last available dietary assessment. HRs correspond to a 90^th^ to 10^th^ percentile difference in EDISEA within UK Biobank. Outcomes are grouped by ICD-10 chapters (shaded bands) and include mortality and incident disease endpoints; event dates were derived from linkage to national death registries and routinely collected health records. Points indicate HRs and horizontal lines 95% CIs; the vertical dashed line denotes the null (HR=1). Blue points denote associations that remained significant after false discovery rate (FDR) correction, whereas white points denote associations not significant. Models were adjusted for age, sex, ethnicity, education qualification, income, smoking status, alcohol consumption status, and physical activity. For incident disease analysis, participants with the corresponding condition at or before baseline were excluded, and individuals were censored at the earliest of incident event, death, loss to follow-up, or end of available registry linkage.

### Molecular and neuroimaging profiling of the EDISEA

We further investigated the molecular and neuroimaging profiles of EDISEA. In proteome-wide association analysis of 2,920 plasma proteins quantified by Olink platform, we identified 810 proteins significantly associated with EDISEA after FDR correction. The strongest inversely associated proteins included FGF21, CHI3L1, GGT1, OXT and PRSS8, while CEACAM16, DSG2, CNTN1, RGMA, and PTPRS showed positive associations **(Figure 5a)**. We identified 384 candidate mediators that showed significant associations with both EDISEA and mortality after FDR correction, and formally tested their average causal mediation effect (ACME) **(Table S8).** We prioritized 71 proteins showing a mediation proportion >10% for the protein-protein interaction (PPI) analysis. These key mediators formed a densely connected PPI module (**Figure 5b**), comprising inflammatory, endothelial, and tissue-remodelling proteins including ACE2, ICAM1, IL6, IL15, VEGFA, HGF, MMP7, TIMP1/TIMP4, PLAUR/PLAT, REN, and multiple TNF receptor superfamily members. Functional enrichment highlighted pathways related to leukocyte migration/chemotaxis and inflammatory response, with enrichment across cell-surface and extracellular compartments and cytokine/chemokine receptor activity. Top KEGG terms included cytokine-cytokine receptor interaction, viral protein interaction with cytokine and cytokine receptor, rheumatoid arthritis, and lysosome **(Figure 5c)**. EDISEA-associated proteins were broadly concordant with a recent study of proteomic signatures of multiple dietary patterns^29^, converging on immune-inflammatory/cytokine signaling and overlapping key proteins (e.g. ACE2, FGF21, OXT, PLAT/PLAUR, PRSS8, HGF, and TNFRSF family members). While pathway-level architecture was highly consistent, the top mediator candidates differed (e.g. GDF15, FABP4, LEP, STC1, FSTL3 in the previous study, CHI3L1, ACE2, TPP1, HGF, CTSD in ours), likely reflecting differences in dietary exposure definitions and analytical objectives. At the organismal level, higher EDISEA was associated with more favorable protein-based organ-specific aging gaps (OAGs, **Figure 5d**), with the strongest association detected for lung age gap (β per 90^th^ to 10^th^ percentile difference = −0.048, 95% CI -0.082 - -0.015), followed by liver and pancreas age gaps.

**Figure 5.**
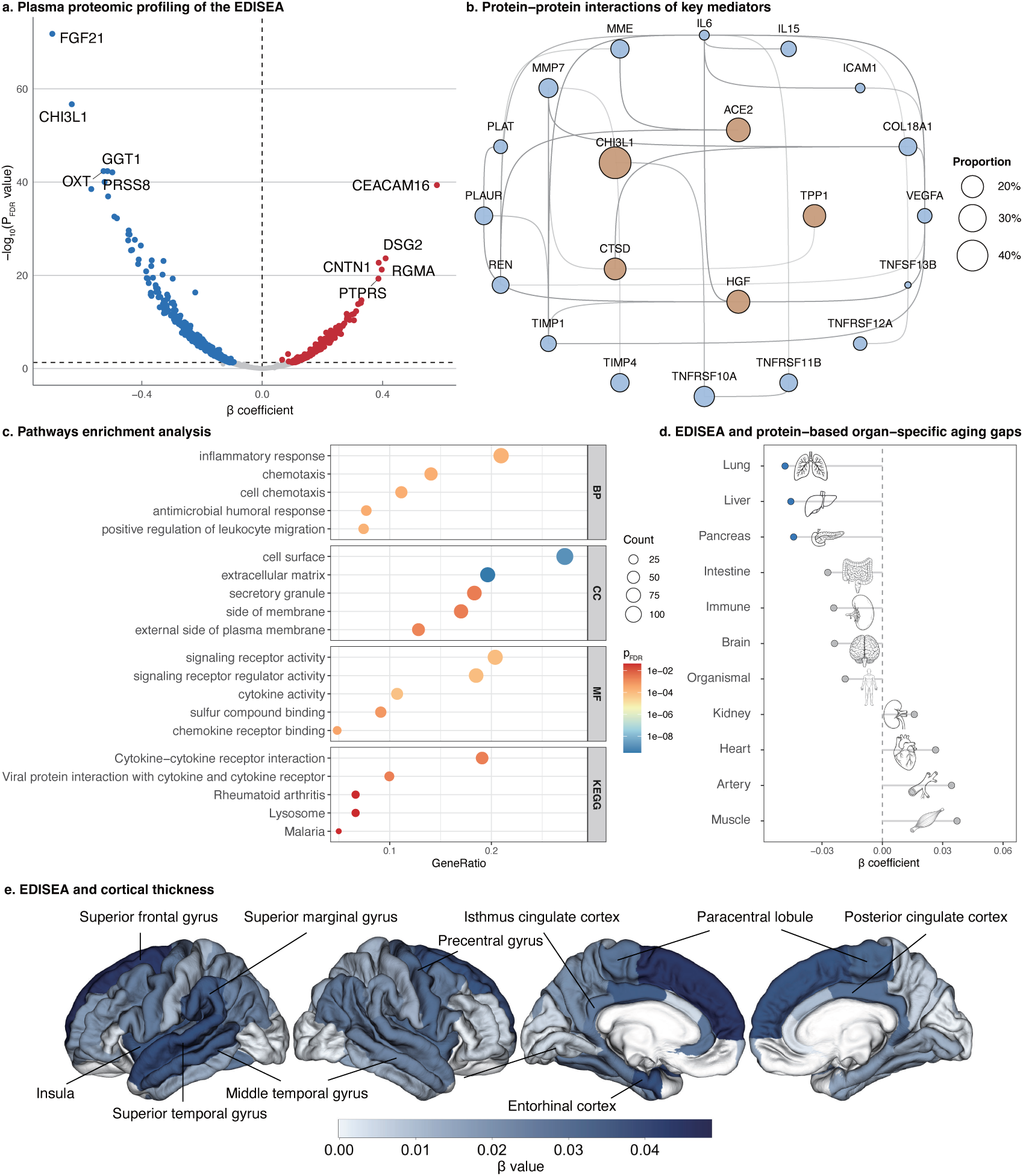
Plasma proteomics, mediation network and neuroimaging associations of EDISEA. **Footnote: a**, Plasma proteomic profiling of EDISEA. Volcano plot of associations between EDISEA and plasma protein abundance (Olink NPX), estimated using multivariable linear regression with each protein (z-scored NPX) as the outcome. The x axis shows the regression coefficient (β) for EDISEA and the y axis shows −log10(FDR-adjusted p). Labeled points denote the most significant proteins. The vertical dashed line marks β=0 and the horizontal dashed line marks the FDR threshold. **b**, Protein-protein interactions of key mediators. Protein-protein interaction (PPI) network for proteins identified as candidate mediators of the EDISEA-mortality association. Edges represent high-confidence interactions from STRING; node size is proportional to the estimated mediation proportion, and highlighted nodes indicate key mediators. **c**, Pathway enrichment analysis. Functional enrichment results for prioritized proteins are shown across Gene Ontology biological process (BP), cellular component (CC), molecular function (MF), and KEGG pathways. Dot size indicates the number of proteins annotated to each term, and color indicates FDR-adjusted p value; the x axis shows GeneRatio. **d**, EDISEA and protein-based organ-specific aging gaps. Associations between EDISEA and organ-specific aging gaps (OAGs) were estimated using multivariable regression and are shown as 90^th^ to 10^th^ difference in EDISEA with 95% confidence intervals. Point color indicates nominal significance (p<0.05 vs ≥0.05). The vertical dashed line denotes β=0. **e**, EDISEA and cortical thickness. Surface maps show cortical regions in which higher EDISEA was associated with thicker cortex, with labeled anatomical regions. The color scale indicates effect size (β) from region-wise regression. UK Biobank covariates included age, sex, ethnicity, education qualification, income, smoking status, alcohol consumption status, physical activity and BMI. Proteomics analysis was additionally adjusted for assessment center, sample measurement batch. Candidate mediators were defined as proteins associated with EDISEA and mortality after FDR correction and with a significant mediation (ACME) after multiple-testing correction; mediation proportion denotes the fraction of the total EDISEA effect statistically explained by the mediator. PPI interactions were derived from STRING and restricted to high-confidence edges; only the largest connected component is displayed. Cortical thickness analysis was additionally adjusted for intracranial volume and assessment center and corrected for multiple comparisons.

Metabolomic profiling **(Figure S4a, S4b)** showed that EDISEA was robustly associated with circulating lipid-related measures and fatty-acid composition, including markers such as total lipoprotein particle concentration and fatty-acid subclass proportions, indicating a coherent metabolic signature. In parallel, EDISEA was associated with a more favorable anti-inflammatory biomarker profile indicated by several inflammation-related biomarkers and composite indices (C-reactive protein, CRP; neutrophils; systemic immune-inflammation index, SII; neutrophil-to-lymphocyte ratio, NLR; platelet-to-lymphocyte ratio, PLR). Altogether, the metabolic and inflammatory pathways partially explained the association between EDISEA and mortality **(Figure S4c)**.

To characterize the neuroimaging associations of EDISEA, we examined cross-sectional associations between EDISEA and 68 regional cortical thickness measures in the UK Biobank brain MRI according to the Desikan-Killiany atlas. Higher EDISEA was associated with greater cortical thickness across 28 out of 68 regions after FDR correction. Strongest associations were observed in left superior frontal gyrus (β per 90^th^ to 10^th^ percentile difference in EDISEA = 0.049, 95% CI 0.027 - 0.071), left superior temporal gyrus (β = 0.042, 95% CI 0.020 - 0.065) and left entorhinal cortex (β = 0.042, 95% CI 0.019 - 0.066). Additional significant regions spanned frontal, temporal and cingulate cortex **(Figure 5e, and Table S9)**, indicating that higher EDISEA was associated with a more favorable cortical thickness profile.

## Discussion

In this multi-cohort study, we developed and externally validated a biologically anchored dietary pattern associated with slower epigenetic aging, termed EDISEA. The EDISEA is characterized by higher consumption of fruits, leafy and yellow vegetables, tea and low-fat dairy products, and lower consumption of processed meats, alcohol (beer and liquor), refined grains, sugar-sweetened beverages and high-fat dairy. Using extensive phenotypic, molecular, and neuroimaging data, we demonstrated that higher adherence to EDISEA was consistently associated with 3-4 years of decelerated epigenetic aging and ∼20% lower mortality risk, as well as a wide range of aging-related outcomes, including incident cancer, dementia, heart disease and stroke. These beneficial associations were partly explained by pathways related to systemic inflammation and immune-cell trafficking, with convergent signals implicating cytokine/chemokine signaling, endothelial activation and vascular growth pathways, and extracellular matrix remodeling. Collectively, these findings support a central role of diet in modulating biological aging and identify a dietary pattern with potential to target epigenetic aging and promote healthy longevity.

EDISEA shares several recommended components with established dietary patterns, including the Alternative Healthy Eating Index (AHEI), Dietary Approaches to Stop Hypertension (DASH), Mediterranean-style diets and the MIND diet^15–18,30,31^, yet it is uniquely designed to comprehensively capture the dietary potential for slowing epigenetic aging. Consistent with these indices, higher adherence to EDISEA emphasizes plant-based, antioxidant-rich foods while minimizing processed foods and high-fat animal products, aligning closely with current dietary guidelines for chronic disease prevention. Prior prospective studies have shown that greater adherence to AHEI or Mediterranean-style diets is associated with nearly a twofold higher likelihood of healthy aging, with key contributors including higher intakes of fruits, vegetables, whole grains and legumes, and lower consumption of red or processed meats, sodium and sugar-sweetened beverages^32^. Building on this evidence, EDISEA reinforces the fundamental role of diet quality in healthy aging while further capturing specific elements highlighted in brain-focused dietary patterns, such as tea consumption and selective alcoholic beverage choices, which are putatively rich in neuroprotective polyphenols and associated with slower cognitive decline and reduced dementia risk^33,34^. While coffee consumption has been linked to favorable cardiometabolic and longevity profiles in several previous investigations^35^, the direction observed here may suggest certain dietary factors may influence epigenetic aging through complex or context-dependent pathways.

In addition, previous empirical data-driven indices focused on intermediate phenotypes, such as the Empirical Dietary Inflammatory Pattern (EDIP) and Empirical Dietary Index for Hyperinsulinemia (EDIH), have shown stronger associations with some health outcomes like cancer than a priori-based diet scores^22,24^. Using similar approaches, our findings indicate that an epigenetic clock anchored diet index can complement and potentially enhance existing dietary metrics when the goal is to predict or modify aging trajectories. Across cohorts, higher EDISEA was consistently associated with a broad spectrum of healthier aging outcomes, including mortality, major chronic diseases, functional limitations and multimorbidity. Outcome-wide analysis in the UK Biobank further supported broad associations across health outcomes, with prominent signals in cardiometabolic, cardiopulmonary diseases, several aging-linked cancers, and end-organ outcomes, suggesting the potential role of EDISEA in preventing multiple aging-related health outcomes.

Omic and imaging-based integrative analyses provide preliminary clues to pathways that may link EDISEA to healthier aging. Proteomic mediation highlighted a connected inflammatory-endothelial and tissue-remodelling module and enrichment for leukocyte migration, chemotaxis and cytokine signaling, consistent with immune-vascular processes contributing to the EDISEA-mortality association^36–38^. These patterns are broadly concordant with recent UK Biobank evidence showing that multiple healthy dietary patterns share reproducible immune-inflammatory/cytokine-related proteomic signatures, suggesting that the immune-vascular signals we observed may reflect shared downstream biology of healthier diets rather than an idiosyncratic feature of any single dietary index^29^. Notably, the specific proteins most strongly implicated in mediation between EDISEA and all-cause mortality only partially overlap (e.g. FGF21, GGT1), implying anchoring diet to epigenetic aging may prioritize a distinct subset of inflammatory-endothelial and tissue-remodelling pathways that are particularly relevant to aging trajectories. We also observed associations with NMR metabolic profiles, supporting links to cardiometabolic regulation^39^. In brain MRI, higher EDISEA was associated with greater cortical thickness across fronto-temporal, insular and cingulate regions, aligning with areas vulnerable to aging-related shrinkage^40^. Together, these signals add biological plausibility to the potential of the EDISEA in slowing epigenetic aging and underscore the need for experimental studies to clarify the exact mechanisms.

These findings have several important implications for both population-level prevention strategies and individual clinical care. From a public health perspective, EDISEA translates complex epigenetic aging biology into a pragmatic, food-based dietary score that aligns closely with existing dietary guidelines. It informs population-wide interventions aimed at slowing biological aging and reducing the burden of multiple aging-related chronic diseases simultaneously. In clinical settings, EDISEA provides a complementary tool for risk stratification. As the EDISEA is currently qualitative with respect to food groups, its further development into a scalable tool for intervention, coupled with validation through randomized trials, could enable clinicians to use the EDISEA principles to guide dietary recommendations for middle-aged and older adults at heightened risk of accelerated aging, cardiometabolic diseases, or neurodegeneration. Future implementation studies will be critical to determine whether improving adherence to EDISEA translates into meaningful clinical benefits, and to identify subgroups most likely to benefit from such interventions.

This study has several limitations that warrant consideration. First, the observational design precludes definitive causal inference. Although we adjusted for a wide range of demographic, socioeconomic, lifestyle, and clinical covariates, residual confounding by unmeasured or imperfectly measured health behaviours and correlated socioeconomic advantages cannot be fully excluded. Secondly, dietary intake was assessed by self-report, which introduces measurement error and recall biases, although misclassification of dietary exposure is likely to be largely non-differential with respect to the outcomes and tend to bias the observed associations toward the null. Dietary assessment methods also differed across cohorts, and while harmonization was carefully performed, some residual heterogeneity is unavoidable. Nonetheless, harmonization and energy adjustment help mitigate some error. Future studies might incorporate objective dietary biomarkers to validate and refine EDISEA components. Third, the generalizability of the findings to more diverse ethnic and cultural settings warrants future investigation as most participants were of European ancestry. In addition, EDISEA was developed and validated primarily in older population, the applicability to younger populations remains to be established. Fourth, this study focused on GrimAge, one of the most well-validated DNA methylation-based clocks for predicting morbidity and mortality. Nevertheless, GrimAge does not capture all biological processes underlying epigenetic aging. It remains possible that dietary exposures influence methylation at CpG sites not incorporated. Although EDISEA showed moderate correlations with several other epigenetic clocks, further investigation using a broader range of epigenetic aging measures is warranted. Finally, our proteomic and neuroimaging analyses are exploratory and cross-sectional, and future studies are needed to replicate these findings and experimentally interrogate the pathways.

### Conclusion

In conclusion, this study developed and validated a data-driven dietary index anchored to epigenetic aging, named EDISEA, and found that greater adherence is consistently associated with slower epigenetic aging and more favorable aging-related outcomes across cohorts. Multi-omic analysis lends credence to the notion that improving diet quality is associated with deceleration in biological aging pathways. These findings support the value of biologically anchored dietary patterns for studying, and potentially informing nutrition strategies for healthy aging, while motivating further longitudinal and interventional validation.

## Methods

### Study design and participants

We conducted a multi-cohort investigation drawing on four large, population-based datasets with complementary designs and populations to develop and evaluate the Empirical Dietary Index for Slower Epigenetic Aging (EDISEA). We derived EDISEA in the U.S. Health and Retirement Study (HRS, specified as the developmental cohort), a longitudinal cohort of older adults. External validation of its correlations with epigenetic clocks was then performed in an independent U.S. sample from the National Health and Nutrition Examination Survey (NHANES). We further examined associations between EDISEA and aging-related health outcomes in the HRS and English Longitudinal Study of Ageing (ELSA). Finally, we performed broad exploratory analyses in the UK Biobank, integrating outcome-wide association analyses, proteomic, metabolic, and neuroimaging assessments.

HRS is a nationally representative longitudinal cohort of adults over age 50 years in the U.S., fielded biennially since 1992 and comprising more than 37,000 individuals in 23,000 households^41^. To maintain representativeness of the U.S. population over age 50, HRS refreshes the sample by enrolling new cohorts of adults in their early fifties and their spouses at regular intervals. In this study, we used HRS data harmonized by the RAND Corporation and linked these survey data to biomarker and epigenetic data derived from biospecimens collected in the HRS enhanced face-to-face assessments. The HRS was approved by the University of Michigan.

NHANES is a nationally representative, repeated, cross-sectional U.S. study of adults across the age spectrum (https://www.cdc.gov/nchs/nhanes/index.htm). Each cycle includes an in-home interview followed by standardized physical examinations and biospecimen collection in Mobile Examination Centers. For prospective analyses, vital status and cause of death are ascertained through linkage to the National Death Index via the NCHS Linked Mortality Files, enabling long-term mortality follow-up. We used data from ten two-year cycles of continuous NHANES (1999-2000 through 2017-2018) and harmonized variables across cycles according to the NHANES documentation. When pooling multiple two-year cycles, we accounted for the NHANES complex survey design by incorporating the appropriate survey weights, strata and primary sampling units, and rescaled the component-specific weights to reflect the combined analytic period, consistent with NHANES analytic guidance. Use of NHANES was approved by the Institutional Review Board of the National Center for Health Statistics.

ELSA is a nationally representative panel study of men and women living in England over age 50 years, initiated in 2002 with an original sample of 11,391 participants spanning ages 50-100 years and followed up biennially^42^. Data are collected using computer-assisted personal interviews and self-completion questionnaires. We used data from wave 9 (2018-2019) to wave 11 (2023-2024). The ELSA was approved by the National Research Ethics Service (London Multicentre Research Ethics Committee).

The UK Biobank is a prospective cohort study of ∼500,000 adults aged 40-69 years recruited in the United Kingdom in 2006-2010^43^. At baseline, participants completed extensive touchscreen questionnaires and interviews, underwent physical measurements, and provided biological samples; repeat assessments and enhanced online assessments (including web-based dietary instruments in subsets) were conducted during follow-up. Longitudinal health outcomes are ascertained through linkage to routinely collected electronic health records, including hospital admissions, cancer registrations, and national death registries, supporting follow-up for incident diseases and mortality. Use of the UK Biobank was approved by the North West Multi-Center Research Ethics Committee. The current analyses were performed under application 816009.

Participants in all cohorts provided informed consent, and each study was approved by local ethics committees.

### Assessment of diet

In HRS, diet was assessed using the validated semi-quantitative food frequency questionnaire (FFQ)^44^ administered in the 2013 Health Care and Nutrition Study (HCNS), an off-year mail survey linked to the main HRS cohort. The FFQ captured respondents’ habitual consumption of a wide range of foods and beverages over the preceding year, with participants reporting intake frequencies for standard portion sizes. From 8,035 participants who completed the questionnaire, we excluded 625 individuals with implausible total energy intake (<500 or >3,500 kcal/day for women and <800 or >4,200 kcal/day for men) and 12 individuals with missing outcome information, yielding a final analytic sample of 7,398 participants.

In NHANES, participants completed at least one interviewer-administered 24-hour dietary recall using the USDA Automated Multiple-Pass Method (AMPM), a standardized, computerized method for dietary recall^45^. The first recall was conducted in person at the Mobile Examination Center, and a second recall (for most participants) was obtained by telephone on a non-consecutive day using the same AMPM protocol. For the present analyses, when two-day 24-hour dietary recalls were available, intakes were averaged across days to approximate usual intake (87.5% of participants, among survey cycles with two-day recalls). From 89,443 participants with completed questionnaires, we excluded 5,973 participants with implausible total energy intake using the same criteria described above. We restricted NHANES to participants aged over 45 years who were eligible for mortality linkage and with over 24 months of follow-up, leaving 23,830 participants for analysis.

In ELSA Wave 9 (2018-2019), diet was assessed using the Oxford WebQ, a validated self-administered online 24-h recall that queries up to ∼200 foods and beverages and provides detailed estimates of energy and nutrient intake^46,47^. Prior validation studies have shown nutrient intakes derived from the questionnaire correlate well with nutrient biomarkers assessed from 24-hour urine samples^48^. From 4,960 participants who completed the WebQ, we excluded 65 participants with implausible total energy intake, leaving 4,895 participants for analysis.

In the UK Biobank, 24-hour dietary recalls were collected at recruitment for participants recruited after Sep 2009 using the Oxford WebQ, and on four additional follow-up rounds after baseline through email to all participants. For participants completing more than one recall, the averages of each food group were calculated. From 210,739 participants completing questionnaires on at least one occasion, we first excluded 21,175 participants who self-reported that their daily diet changed on a weekly basis (Field ID 1548) and a further 2,529 participants with implausible total energy intake. After these exclusions, 187,035 UK Biobank participants were retained.

We harmonized and aggregated individual food items into 38 food groups (e.g. dark leafy vegetables, other vegetables, fruits, whole grains, nuts, red meat, processed meat, sugary foods, etc.) in the four cohorts^49^. Nutrient intakes were energy-adjusted using the residual method^50^ to reduce measurement error and confounding by total caloric intake.

### Epigenetic clocks

The primary response variable for EDISEA derivation was GrimAge acceleration (GrimAgeAccel), defined as the residual from regressing DNAm-based GrimAge on chronological age^51^. GrimAge is a second-generation epigenetic clock developed to capture mortality- and morbidity-related biological aging by combining DNAm-based surrogates for multiple plasma proteins together with a DNAm-based estimate that is highly relevant to aging and chronic diseases, yielding a composite that maps onto inflammation, cardiometabolic, and vascular biology, plausibly modifiable by diet^51^. GrimAge has consistently demonstrated superior performance compared with earlier first-generation clocks in predicting time-to-death and major aging-related outcomes^7,52,53^, and is therefore well-suited as a biologically and clinically informative endpoint for constructing a dietary index intended for slower epigenetic aging.

In HRS, whole-blood DNA was profiled on the Illumina Infinium MethylationEPIC BeadChip v1.0 (assayed at University of Minnesota), and DNAm-based epigenetic aging measurements have been derived and analyzed in prior studies^54–56^. Pre-processing and quality control were conducted in R, including probe filtering based on detection P values (threshold 0.01), and removal of low-quality samples, sex-discordant samples, and control/duplicate arrays. GrimAge values derived from the 2016 Venous Blood Study DNAm release are ultimately computed from the post-QC beta matrix by applying the published GrimAge model coefficients to the required CpG subset. Among participants with valid dietary data, 1,711 were eligible for epigenetic age acceleration analysis.

In NHANES, whole-blood DNAm was profiled on the Illumina EPIC BeadChip arrays (assayed at Duke University). Bioinformatic processing started from conversion to methylated/unmethylated signals, followed by color correction and background subtraction using array control probes, outlier removal, and normalization to address probe-type bias via BMIQ. Epigenetic biomarkers including GrimAge and DunedinPoAm have been released for NHANES 1999-2002 and widely used in prior work^57–59^. GrimAge was produced by applying published coefficients to the corresponding CpG set to generate participant-level scores. DNAm profiling was conducted from the 1999-2000 and 2001-2002 cycles using a design that oversampled minoritized ethnic groups. Among participants with valid dietary data, 2,331 were eligible for epigenetic age acceleration analysis.

### Health outcomes

The primary health outcome of the study was mortality, and the secondary outcomes were aging-related diseases and disability, including cancer, dementia, heart disease, stroke, Activities of Daily Living (ADL) disability, Instrumental Activities of Daily Living (IADL) disability and a broader set of chronic diseases^60^. In the time-to-event analysis, follow-up started at the baseline (dietary) assessment date, and participants were censored at the end of outcome linkage/last completed follow-up, loss to follow-up, or mortality, whichever came first.

In HRS, mortality was ascertained using death information collected in follow-up and linkage-based vital status information through May 2023. Hypertension, diabetes, lung disease, heart disease, stroke, cancer and arthritis were defined from biennial self-reported physician diagnosis or proxy reports. Dementia was defined using validated cognitive-status algorithm based on longitudinal cognitive assessments and proxy information^61^. Incident event time was defined as the first report after baseline. ADL/IADL limitations were derived from standard HRS functional questionnaires and incident rate ratio was defined as the number of new-onset difficulty/dependence compared with baseline. To capture longitudinal, multi-system aging trajectories, we analyzed Pace of Aging (PoA), which was obtained from the published HRS PoA phenotype summarizing longitudinal change across nine repeatedly measured parameters (CRP, cystatin-C, HbA1c, diastolic blood pressure, waist circumference, peak flow, tandem balance, grip strength and gait speed) into a dynamic measure of the rate of biological aging^60^.

In NHANES, mortality was defined using the NHANES Linked Mortality File (National Death Index Linkage, https://www.cdc.gov/nchs/linked-data/mortality-files/index.html). Follow-up was calculated from the MEC examination date to date of death, participants were otherwise censored at the end of the public linkage period (December 2019).

In ELSA, cancer incidence was obtained through linkage to national cancer registries. Heart disease, stroke, and other chronic diseases were defined using linked hospital inpatient records and death registries. Dementia was defined algorithmically, following DSM-IV and other clinical criteria in that it hinges on non-transient impairment in two or more cognitive domains applied to ELSA participants (orientation, immediate and delayed memory, verbal fluency and executive function) resulting in functional impairment. Impairment of a cognitive domain was defined as a score of more than 1.5 SD below the mean compared with the population aged 50-80 years with the same education qualification. Functional impairment was defined as needing assistance with one or more activities of daily living (getting into or out of bed, walking across a room, bathing or showering, using the toilet, dressing, cutting food, and eating)^62^. Incident event time was defined as the midpoint imputation between two consecutive interview dates. ADL/IADL limitations were derived from repeated functional questionnaires, and incident rate ratio was defined as the number of new-onset difficulty/dependence compared with baseline.

In UK Biobank, mortality was ascertained via linkage to national death registries, using the recorded date of death. Cancer incidence was obtained through linkage to national cancer registries using the first recorded diagnosis date. For other incident diseases, we used the UK Biobank first occurrence fields (category 1712), which map diagnosis from primary care, hospital inpatient records, death register records, and self-reported medical conditions to 3-character ICD-10 codes and provide, for each participant, the earliest date that code was recorded across any source. Participants were followed from the baseline assessment date until the earliest of incident disease, death, or administrative censoring at the UK Biobank-recommended data-provider censoring dates for the relevant linked dataset, to minimise incomplete follow-up. Specifically, censoring dates (used as guidance) were 31 Aug 2024 for death data in England & Wales and 30 Nov 2024 in Scotland; for cancer registry data, 31 May 2023 in England, 30 Sep 2023 in Scotland (and 31 Dec 2016 in Wales as per UK Biobank’s noted coverage issue); and for hospital inpatient data, 31 Mar 2023 in England, 31 Aug 2022 in Scotland, and 31 May 2022 in Wales. We analysed a predefined panel of 58 health outcomes grouped into 14 major disease categories besides mortality, consistent with prior literature^63^ **(Table S10)**.

### Brain structural imaging

UK Biobank brain MRI was acquired on a dedicated Siemens Skyra 3T system with a 32-channel head coil at imaging centers in Cheadle Manchester, and from 2017 onwards, additionally sites in Newcastle and Reading^64^. We used UK Biobank’s centrally processed imaging pipeline outputs. Briefly, the T1 pipeline includes gradient distortion correction, brain extraction and nonlinear registration to MNI space, followed by tissue segmentation and additional morphometric outputs. Cortical surface reconstruction was performed with FreeSurfer, using T1 and when available T2-FLAIR to improve cortical modelling. FreeSurfer outputs underwent automated QC using Qoala-T. Details of image processing and quality control can be found in a previous article^65^. For the current analyses, we extracted cortical thickness IDPs from 68 regions of Desikan-Killiany parcellation^66^ (category 192) and tested cross-sectional associations between EDISEA and regional thickness using multivariable regression adjusting for core sociodemographic and lifestyle covariates, and imaging-specific confounds including intracranial volume and assessment center from 39,507 participants.

### Olink plasma proteomic measurement

Plasma proteomics were obtained from the UK Biobank Pharma Proteomics Project (UKB-PPP), which profiled ∼54k participants using the Olink Explore 3072 Proximity Extension Assay (PEA), yielding 2,941 protein analytes (2,923 unique proteins) reported as NPX values (category 1838)^67^. NPX generation and normalization followed Olink/UKB procedures including normalization to extension controls, plate controls, and batch-level alignment. UKB-PPP QC applied a standardized multi-step protocol: removal of Olink control samples, withdrawn or unprocessed participants, detection and removal of outlier samples, filtering observations with QC/assay warnings, and identification of likely sample swaps^68^. We further excluded proteins with values below the lower limits of detection for more than 25% of samples^69^, leaving 2,920 proteins for analysis. For downstream association analyses, proteins were analyzed as z-standardized NPX values and models additionally adjusted for key technical covariates including assessment center and sample measurement batch.

### Organ-specific age gaps

We reproduced a published protein-based organ aging framework^70^ in UK Biobank using Olink plasma proteomics to capture inter-individual deviations in organ-level biological aging, providing mechanistically interpretable intermediate phenotypes linking EDISEA to downstream morbidity and mortality. For each organ, we trained a LightGBM regression model to predict chronological age from corresponding protein panel. Details of proteins with annotation of organ enrichment were available in this article. To obtain unbiased predicted ages for all participants, we generated out-of-fold predictions using 5-fold cross-fitting with early stopping. Organ-specific age gaps were then defined as the residuals from regressing predicted organ age on chronological age, such that positive OAG indicates an older proteomic age than expected for a participant’s chronological age.

### Inflammation markers and metabolites in plasma

Inflammation markers were derived from UK Biobank baseline blood assays. High-sensitivity C-reactive protein (CRP) was measured in serum using an immunoturbidimetric method on the Beckman Coulter AU5800 platform as part of the UK Biobank biomarker panel. When blood-cell-based inflammatory indices were considered (e.g., neutrophil/lymphocyte or platelet components), cell counts and differentials were taken from the UK Biobank haematology measurements (full blood count and differential outputs). Plasma metabolomic profiling used the UK Biobank Nightingale Health NMR metabolic biomarker data (category 220), generated from randomly selected EDTA plasma aliquots and released in multiple phases. The Nightingale NMR platform quantifies 249 metabolic measures per sample, including 168 absolute concentration measures and 81 ratio measures. These data provide broad coverage of circulating lipids, lipoprotein measures, fatty acids and small molecules (e.g., amino acids, ketones, and glycolysis-related metabolites)^71^. Details of sample processing and quality control can be found at https://biobank.ndph.ox.ac.uk/showcase/label.cgi?id=220.

### Assessment of covariates

In this study, we harmonized multiple covariates for confounding adjustment according to previous studies^20^. In the four analytic cohorts, we used questionnaires at study recruitment (NHANES and UK Biobank) or dietary assessment (HRS and ELSA) to collect information on demographics, socioeconomic factors, lifestyles and clinical history, including age, gender, ethnicity (white, others), educational attainment (below college, college and above), income (low, medium, high), smoking status (never smoker, former smoker, current smoker), alcohol consumption status (never drinker, former drinker, current drinker), physical activity (low, medium, high). Baseline body mass index (BMI) was calculated by dividing weight in kilograms by height in meters squared. Baseline clinical history, including hypertension, diabetes, and cerebrovascular diseases was obtained in each cohort. Missing covariates were imputed using multiple imputation by chained equations, with 5 imputed datasets and 5 iterations^72^ **(Table S11)**.

### Statistical analysis

Baseline characteristics of participants in 4 cohorts were summarized as counts with percentages for categorical variables, and means with standard deviations (SDs) for continuous variables.

We developed the EDISEA index in the HRS derivation sample using a data-driven approach by integrating food frequency questionnaires (FFQ)-derived food-group intakes with DNAm-based epigenetic age measures. For each epigenetic clock, epigenetic age acceleration (EAA) was computed as the residual from a linear regression of DNAm-based epigenetic age on chronological age, which was interpreted as years older or younger in epigenetic age. The primary target for EDISEA derivation was GrimAge acceleration. In sensitivity analyses, we additionally generated EAA measures for another second-generation clock (DunedinPoAm^73^).

We first assessed the associations of the 38 food groups with GrimAge acceleration, adjusted for the above-mentioned covariates. For interpretability, each food-group exposure was rescaled to represent a 90^th^ to 10^th^ percentile contrast and modeled in linear regression with GrimAgeAccel. Non-linear tests were performed using restricted cubic splines. Second, the HRS derivation sample was randomly split into 70% training and 30% testing sets for EDISEA construction and internal validation. Within the training set, we fitted elastic-net regression models with GrimAgeAccel as the response variable and 38 food groups as predictors, each z- standardized in the entire cohort. For hyperparameter tuning, we selected mixing parameter α and penalty parameter λ yielding the minimum cross-validated mean squared error using 10-fold cross-validation on the training set. The penalized model in the full training sample was refitted with the selected hyperparameters, and non-zero food-group coefficients were extracted. EDISEA was then computed for each participant as a weighted reversed sum of the standardized selected food groups to predict lower GrimAgeAccel. Internal validation was evaluated in the held-out testing set by computing the Spearman correlation between EDISEA and GrimAgeAccel.

To verify the robustness and generalizability of EDISEA, independent external validation was performed in NHANES. Food groups were z-standardized within NHANES, and HRS-derived EDISEA weights were applied to compute EDISEA. The Spearman correlation between EDISEA and GrimAgeAccel and monotonic patterns across EDISEA quintiles for GrimAgeAccel and other EAA measures were evaluated descriptively.

Multivariable Cox proportional hazard regression models were employed to estimate the associations of EDISEA with mortality and incident aging-related diseases, with results presented as hazard ratio (HR) and 95% confidence interval (CI). Incidence rate ratio (IRR) of count-type outcomes reflecting functional impairment and overall chronic disease burden were estimated using log-linked Poisson regression with an offset for follow-up time to model incidence rates. For incident aging-related diseases, models were adjusted for age, sex, ethnicity, income, education qualification, smoking status, drinking status, physical activity, and BMI. For mortality, models were additionally adjusted for history of hypertension, diabetes and cerebrovascular diseases. Cohort-specific HR and IRR estimates and their standard errors were pooled using inverse-variance weighted random-effects meta-analysis to yield pooled estimates. We evaluated the association between EDISEA and PoA using restricted cubic splines (RCS) within multivariable regression. The number of knots (k) was chosen by minimizing AIC across models with 3-7 knots.

In the UK Biobank, we analyzed a panel of 58 health outcomes grouped into 14 major disease categories in addition to all-cause mortality, according to a previous study^63^. For each outcome, participants with prevalent disease at baseline were excluded from the risk set for the corresponding outcome. Associations between EDISEA and time-to-event outcomes were estimated using Cox proportional hazards regression, with follow-up time defined as time from last dietary recall to event/death/censoring. EDISEA was modelled as a continuous variable, and estimates are reported as hazard ratios with 95% confidence intervals. To account for multiplicity, we controlled the false discovery rate across outcomes. Outcome-specific results were additionally summarized at the disease-category level to facilitate interpretation. We evaluated plasma proteomic signatures of EDISEA and explored proteomic mediation of EDISEA-mortality associations in the subset of participants with Olink plasma proteomics (NPX) data. Each protein NPX value was z-standardized in the analytic sample, and proteome-wide associations with EDISEA were estimated using multiple linear regression models. To evaluate whether EDISEA-related proteins may statistically mediate the association between EDISEA and mortality, we implemented causal mediation analysis using quasi-Bayesian approximation, followed by over-representation analysis for Gene Ontology (GO) and KEGG pathways. Specifically, for each protein we first fitted a linear regression mediator model to estimate the EDISEA-protein association, and then fitted a parametric survival regression model with both EDISEA and the protein included. We then used the *mediate* function in the R package mediation^74^ to estimate the average causal mediation effect (ACME) with uncertainty quantified via the quasi-Bayesian simulation approach. Proteins with a mediation proportion >10% were kept in protein-protein interaction analysis utilizing STRING software^75^. Associations between EDISEA and metabolites and inflammation markers in blood were estimated with the same analysis. Structural equation modelling^76^ was employed to estimate direction-dependent relationships between EDISEA and mortality mediated by inflammation and metabolism.

All analyses were performed in R 4.5.0 (http://www.r-project.org/), with two-sided p<0.05 as the threshold for statistical significance. Multiple testing was addressed using the Benjamini-Hochberg false discovery rate (FDR) correction, applied for each hypothesis family.

## Data availability

The data used in this study are available from the respective cohorts under data access agreements and restrictions. The Health and Retirement Study (HRS) data are available through the University of Michigan and the National Archive of Computerized Data on Aging (NACDA) subject to their data use policies. NHANES data are publicly available from the U.S. Centers for Disease Control and Prevention. English Longitudinal Study of Ageing (ELSA) data are available via the UK Data Service under standard access procedures. The UK Biobank data are available through application to the UK Biobank (application ID: 816009).

## Acknowledgements

We thank the participants and staff of HRS, NHANES, ELSA, and UK Biobank for their invaluable contributions. This study was supported by the Fundamental Research Funds for the Central Universities (2025ZFJH01, to BL). The funders had no role in the study design or implementation; data collection, management, analysis or interpretation; manuscript preparation, review or approval; the decision to publish.

## Author contributions

SL, LZ, CY, HC, and BL planned and designed the study. SL and LZ accessed and verified the data, did the statistical analysis and drafted the article. SL, LZ, and HC checked the accuracy of the statistical analysis. BL, HC, CY, JY, MW, GP, GZ, HM, and WC contributed to reviewing and editing. All authors participated in the interpretation of the results and critical revision of the article. All authors had full access to all the data in the study and accept responsibility for the decision to submit for publication. BL is the guarantor of this work, and has final responsibility for the integrity of the data, the accuracy of the data analysis and the decision to submit for publication.

