## Supplementary materials for "A data-driven dietary pattern anchored to slower epigenetic aging is associated with a spectrum of aging-related health outcomes"

### Supplementary Figures

**Figure S1. Study population inclusion flow chart**

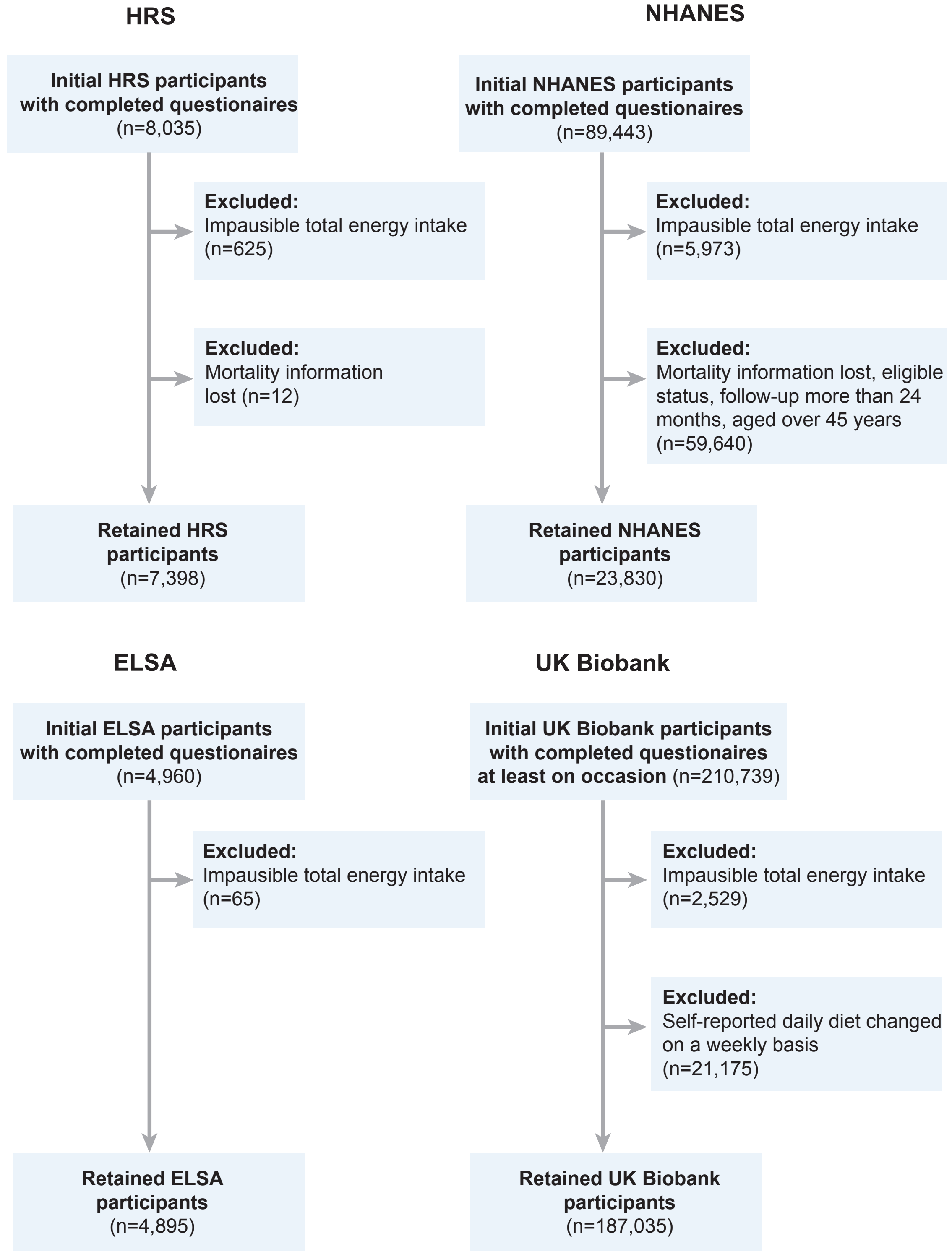

**Footnote:** Flow diagram summarizing inclusion and exclusion of participants from HRS, NHANES, ELSA, and UK Biobank. Numbers indicate unique participants with completed baseline dietary assessment and subsequent exclusions applied sequentially to derive the final analytic samples used in primary analysis. Implausible total energy intake denotes participants with implausible reported daily caloric intake based on pre-specified cutoffs/QC rules (see Methods).

**Figure S2. Correlations between different dietary patterns**

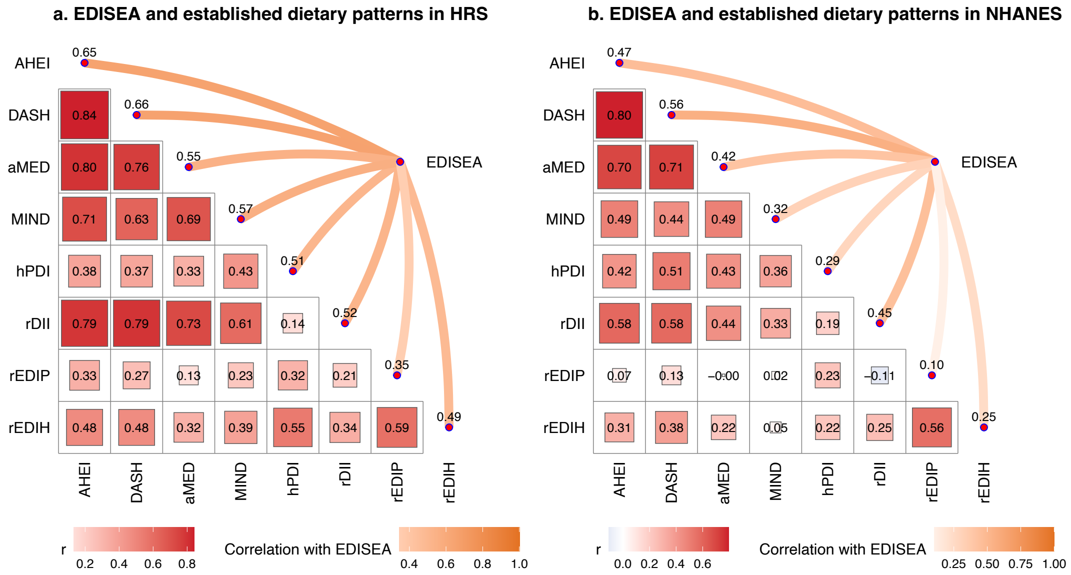

**Footnote:** Correlation matrices display pairwise Spearman correlations among dietary pattern scores within HRS (**a**) and NHANES (**b**). Curved links denote correlations between EDISEA and each established dietary pattern score; link color intensity corresponds to the magnitude of correlation with EDISEA. Square tiles represent pairwise correlations between established dietary patterns. Correlations were computed in subcohorts with epigenetic age data available. DII, EDIH and EDIP were reversed to rDII, rEDIH, rEDIP respectively so that higher values always reflect healthier dietary patterns. AHEI, Alternate Healthy Eating Index; DASH, Dietary Approaches to Stop Hypertension; aMED, alternate Mediterranean diet score; DII, Dietary Inflammatory Index; hPDI, healthy Plant-based Diet Index; MIND, Mediterranean-DASH Intervention for Neurodegenerative Delay; EDIH/EDIP, empirical dietary indices for hyperinsulinemia/inflammatory potential.

**Figure S3. Association between EDISEA and Pace of Aging (PoA) in HRS**

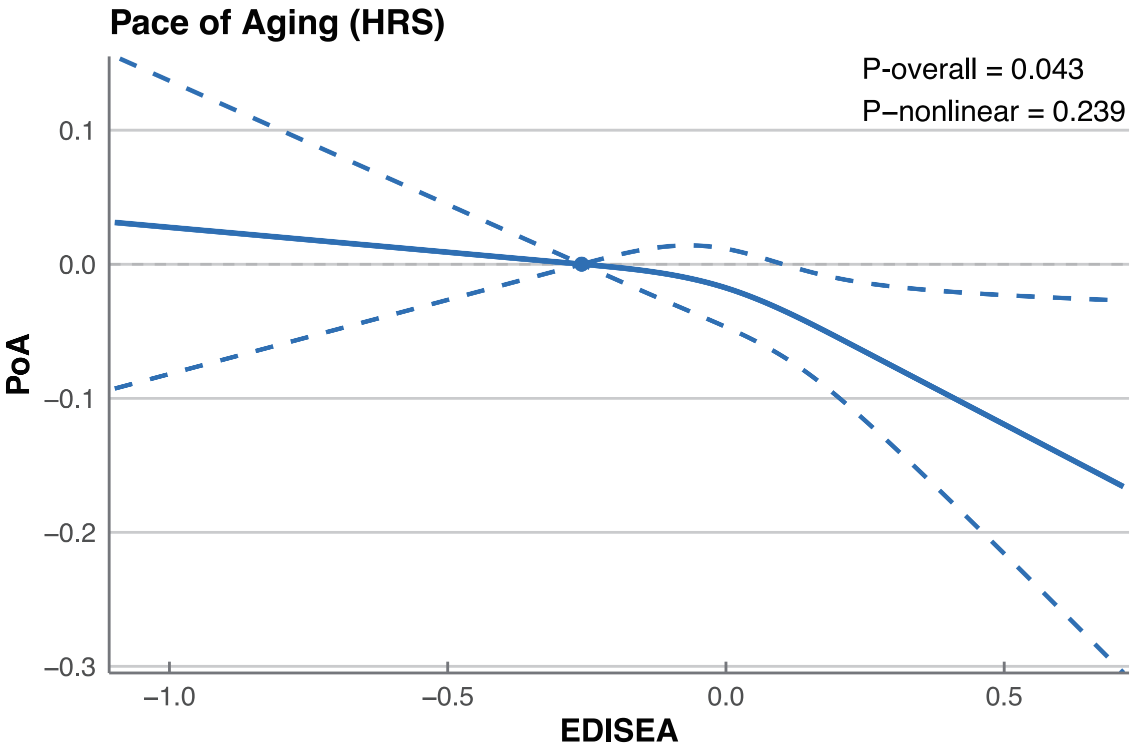

**Footnote:** Restricted cubic spline model showing the multivariable-adjusted association between continuous EDISEA and Pace of Aging (PoA) in HRS. Solid line indicates the adjusted estimate and dotted line the 95% confidence interval. The reference value for EDISEA was set at the 10$^{th}$ percentile. Overall and nonlinearity P values are shown (P-overall = 0.043, P-nonlinear = 0.239).

**Figure S4. Metabolism and inflammation profiling in UK Biobank**

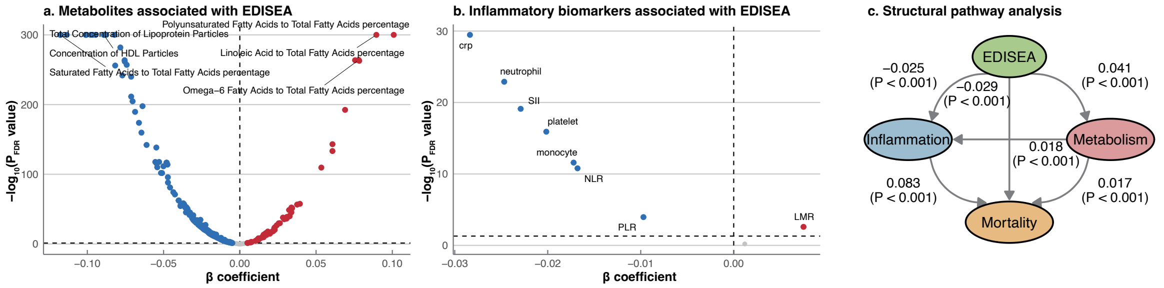

**Footnote: a**, Volcano plot of circulating metabolites associated with EDISEA. Each point represents one metabolite; the x-axis shows the multivariable-adjusted regression coefficient (β) for EDISEA-metabolite association and the y-axis shows -log10(FDR-adjusted p value). **b**, Volcano plot of inflammatory biomarkers associated with EDISEA using the same modeling framework and visualization. **c**, Structural pathway model summarizing direct and indirect associations linking EDISEA, metabolomic and inflammatory signatures, and mortality; numbers on paths denote standardized effect estimates with corresponding p values. CRP, C-reactive protein; SII, systemic immune-inflammation index; NLR, neutrophil-to-lymphocyte ratio; PLR, platelet-to-lymphocyte ratio; LMR, lymphocyte-to-monocyte ratio.

### Supplementary Tables

### Table S1. Characteristics of the study population at baseline

| Characteristics | HRS<br>N = 7,398 | NHANES<br>N = 23,830 | ELSA<br>N = 4,895 | UK Biobank<br>N = 187,035 |
|---|---|---|---|---|
| **Age, years, mean (SD)** | 68.7 (11.0) | 62.7 (11.2) | 68.0 (7.9) | 56.1 (8.0) |
| **Total energy intake (kcal/d), mean(SD)** | 1,784.8 (673.5) | 1,875.9 (679.9) | 1,993.7 (548.3) | 2,045.9 (549.9) |
| **Sex, n (%)** | | | | |
| Male | 3,022 (41%) | 11,557 (48%) | 2,206 (45%) | 83,956 (45%) |
| Female | 4,376 (59%) | 12,273 (52%) | 2,689 (55%) | 103,079 (55%) |
| **Ethnicity, n (%)** | | | | |
| White | 5,691 (77%) | 11,924 (50%) | 4,755 (97%) | 179,579 (96%) |
| Others | 1,707 (23%) | 11,906 (50%) | 140 (2.9%) | 7,456 (4.0%) |
| **Education, n (%)** | | | | |
| Below college | 3,674 (50%) | 18,912 (79%) | 3,503 (72%) | 106,917 (57%) |
| College and above | 3,724 (50%) | 4,918 (21%) | 1,392 (28%) | 80,118 (43%) |
| **Income, n (%)** | | | | |
| Low | 2,099 (28%) | 6,665 (28%) | 1,805 (37%) | 102,722 (55%) |
| Medium | 1,952 (26%) | 9,207 (39%) | 2,198 (45%) | 59,992 (32%) |
| High | 3,347 (45%) | 7,958 (33%) | 892 (18%) | 24,321 (13%) |
| **Smoking status, n (%)** | | | | |
| Never | 3,360 (45%) | 11,701 (49%) | 2,006 (41%) | 106,544 (57%) |
| Former smoker | 3,240 (44%) | 7,970 (33%) | 2,563 (52%) | 66,369 (35%) |
| Current smoker | 798 (11%) | 4,159 (17%) | 326 (6.7%) | 14,122 (7.6%) |
| **Alcohol consumption, n (%)** | | | | |
| Never | 3,356 (45%) | 3,771 (16%) | 177 (3.6%) | 6,117 (3.3%) |
| Former | 1,163 (16%) | 4,075 (17%) | 352 (7.2%) | 5,717 (3.1%) |
| Current | 2,879 (39%) | 15,984 (67%) | 4,366 (89%) | 175,201 (94%) |
| **Physical activity, n (%)** | | | | |
| Low | 2,390 (32%) | 14,983 (63%) | 3,129 (64%) | 33,999 (18%) |
| Medium | 2,673 (36%) | 5,435 (23%) | 522 (11%) | 79,361 (42%) |
| High | 2,335 (32%) | 3,412 (14%) | 1,244 (25%) | 73,675 (39%) |
| **BMI (kg/m$^2$), n (%)** | | | | |
| 0-24.9 | 2,068 (28%) | 6,128 (26%) | 1,431 (29%) | 71,521 (38%) |
| 25-29.9 | 2,743 (37%) | 8,620 (36%) | 2,005 (41%) | 77,556 (41%) |
| >=30 | 2,587 (35%) | 9,082 (38%) | 1,459 (30%) | 37,958 (20%) |
| **History of health conditions, n (%)** | | | | |
| Diabetes | 1,755 (24%) | 4,947 (21%) | 557 (11%) | 7,674 (4.1%) |
| Hypertension | 4,474 (60%) | 16,988 (71%) | 2,053 (42%) | 45,266 (24%) |

| | | | | |
|---|---|---|---|---|
| Cardiovascular diseases | 2,222 (30%) | 4,094 (17%) | 1,173 (24%) | 6,204 (3.3%) |

**Footnote:** Values are presented as mean (SD) for continuous variables and n (%) for categorical variables. Sample sizes reflect participants with non-missing EDISEA and required outcome linkage in each cohort. Total energy intake represents average daily caloric intake derived from cohort-specific dietary instruments and harmonized to kcal/day. Ethnicity was harmonized as White vs Other based on cohort-specific ethnicity categories. Education was harmonized as below college vs college and above. Household income was categorized into low/medium/high within each cohort using cohort-specific income measures. Smoking status and alcohol consumption status were harmonized as never/former/current. Physical activity was harmonized into low/medium/high using cohort-specific activity questionnaires/metrics. BMI categories were defined as <25.0, 25.0-29.9, and ≥30.0 kg/m². History of diabetes, hypertension, and cardiovascular diseases indicates self-reported or clinically ascertained physician diagnosis at baseline, harmonized across cohorts.

**Table S2. Elastic-net selected food groups for predicting GrimAge acceleration and stability across cross-validation folds and penalty parameters**

| Main model | | Cross-validation stability | | | | Penalty parameter stability | | | |
|---|---|---|---|---|---|---|---|---|---|
| Food groups | Estimate | Food groups | Selection frequency | Estimate median | Estimate IQR | Food groups | Selection frequency | Estimate median | Estimate IQR |
| Beer | 0.097 | Beer | 1 | 0.145 | 0.019 | Beer | 1 | 0.107 | 0.007 |
| Processed meat | 0.082 | Processed meat | 1 | 0.122 | 0.017 | Processed meat | 1 | 0.095 | 0.006 |
| Sugar-sweetened beverages | 0.073 | Liquor | 1 | 0.089 | 0.012 | Green leafy vegetables | 1 | -0.063 | 0.003 |
| Fruits | -0.056 | Green leafy vegetables | 1 | -0.085 | 0.011 | Tea | 1 | -0.054 | 0.003 |
| Liquor | 0.042 | Tea | 1 | -0.085 | 0.015 | Liquor | 1 | 0.051 | 0.002 |
| Tea | -0.035 | Sugar-sweetened beverages | 1 | 0.075 | 0.011 | Fruits | 1 | -0.050 | 0.000 |
| Green leafy vegetables | -0.034 | Refined grains | 1 | 0.063 | 0.018 | Refined grains | 1 | 0.035 | 0.002 |
| Refined grains | 0.030 | Coffee | 1 | 0.049 | 0.022 | Sugar-sweetened beverages | 1 | 0.033 | 0.002 |

| | | | | | | | | | |
|---|---|---|---|---|---|---|---|---|---|
| Dark yellow vegetables | -0.016 | Fruits | 1 | -0.048 | 0.008 | Dark yellow vegetables | 1 | -0.024 | 0.002 |
| Coffee | 0.015 | Organ meat | 1 | 0.044 | 0.010 | Coffee | 1 | 0.010 | 0.001 |
| High-fat dairy | 0.013 | High-fat dairy | 1 | 0.041 | 0.014 | Fries | 1 | 0.002 | 0.001 |
| Low-fat dairy | -0.006 | Dark yellow vegetables | 1 | -0.035 | 0.018 | High-fat dairy | 1 | 0.002 | 0.000 |
| Butter | 0 | Oils | 1 | -0.029 | 0.013 | Artificial sweetened beverages | 0 | 0 | 0 |
| Margarine | 0 | Pizza | 1 | -0.021 | 0.020 | Butter | 0 | 0 | 0 |
| Fruit juice | 0 | Fries | 0.9 | 0.038 | 0.010 | Condiments | 0 | 0 | 0 |
| Tomato | 0 | Low-fat dairy | 0.9 | -0.013 | 0.005 | Cruciferous vegetables | 0 | 0 | 0 |
| Cruciferous vegetables | 0 | Fruit juice | 0.8 | -0.012 | 0.009 | Eggs | 0 | 0 | 0 |
| Legumes | 0 | Mayonnaise | 0.8 | 0.009 | 0.023 | Fish and other seafoods | 0 | 0 | 0 |
| Other vegetables | 0 | Other vegetables | 0.7 | 0.027 | 0.035 | Fruit juice | 0 | 0 | 0 |
| Eggs | 0 | Margarine | 0.7 | 0.008 | 0.014 | Garlic | 0 | 0 | 0 |
| Red meat | 0 | Wine | 0.7 | -0.003 | 0.010 | Legumes | 0 | 0 | 0 |
| Poultry | 0 | Snack | 0.6 | -0.010 | 0.024 | Low-fat dairy | 0 | 0 | 0 |
| Organ meat | 0 | Legumes | 0.6 | 0.004 | 0.009 | Margarine | 0 | 0 | 0 |

| | | | | | | | | | |
|---|---|---|---|---|---|---|---|---|---|
| Fish and other seafoods | 0 | Garlic | 0.5 | 0.001 | 0.008 | Mayonnaise | 0 | 0 | 0 |
| Whole grains | 0 | Tomato | 0.5 | 0.000 | 0.002 | Nuts | 0 | 0 | 0 |
| Pizza | 0 | Butter | 0.4 | 0 | 0.004 | Oils | 0 | 0 | 0 |
| Potato | 0 | Cruciferous vegetables | 0.4 | 0 | 0.006 | Organ meat | 0 | 0 | 0 |
| Fries | 0 | Nuts | 0.2 | 0 | 0 | Other vegetables | 0 | 0 | 0 |
| Snack | 0 | Artificial sweetened beverages | 0.1 | 0 | 0 | Pizza | 0 | 0 | 0 |
| Artificial sweetened beverages | 0 | Condiments | 0.1 | 0 | 0 | Potato | 0 | 0 | 0 |
| Wine | 0 | Eggs | 0.1 | 0 | 0 | Poultry | 0 | 0 | 0 |
| Sweets | 0 | Potato | 0.1 | 0 | 0 | Red meat | 0 | 0 | 0 |
| Nuts | 0 | Poultry | 0.1 | 0 | 0 | Snack | 0 | 0 | 0 |
| Soup | 0 | Red meat | 0.1 | 0 | 0 | Soup | 0 | 0 | 0 |
| Condiments | 0 | Soup | 0.1 | 0 | 0 | Sweets | 0 | 0 | 0 |
| Garlic | 0 | Sweets | 0.1 | 0 | 0 | Tomato | 0 | 0 | 0 |
| Mayonnaise | 0 | Whole grains | 0.1 | 0 | 0 | Whole grains | 0 | 0 | 0 |
| Oils | 0 | Fish and other seafoods | 0 | 0 | 0 | Wine | 0 | 0 | 0 |

**Footnote:** The elastic-net model was used to identify food groups jointly associated with GrimAge acceleration and to derive EDISEA component weights. Main model reports the fitted elastic-net coefficients from the primary specification. Cross-validation stability summarizes refits across cross-validation folds using the primary penalty selection rule. Selection frequency is the proportion of folds in which a food group's coefficient was non-zero. Estimate median and estimate IQR are the median and interquartile range of the non-zero coefficients across folds. Penalty parameter stability evaluates robustness to the choice of the regularization strength (λ) within the prespecified search grid. Selection frequency, median coefficient, and IQR are computed analogously across λ refits.

**Table S3. Elastic-net selected food groups for predicting DunedinPoAm, and stability across cross-validation folds and penalty parameters**

| Main model | | Cross-validation stability | | | | Penalty parameter stability | | | |
|---|---|---|---|---|---|---|---|---|---|
| Food groups | Estimate | Food groups | Selection frequency | Estimate median | Estimate IQR | Food groups | Selection frequency | Estimate median | Estimate IQR |
| Sugar-sweetened beverages | 0.074 | Green leafy vegetables | 1 | -0.076 | 0.020 | Green leafy vegetables | 1 | -0.071 | 0.004 |
| Liquor | 0.067 | Liquor | 1 | 0.065 | 0.013 | Liquor | 1 | 0.058 | 0.003 |
| Fruits | -0.047 | Processed meat | 1 | 0.047 | 0.024 | Processed meat | 1 | 0.048 | 0.002 |
| Processed meat | 0.045 | Sugar-sweetened beverages | 1 | 0.040 | 0.015 | Sugar-sweetened beverages | 1 | 0.038 | 0.001 |
| Tea | -0.027 | Organ meat | 1 | 0.040 | 0.009 | Organ meat | 1 | 0.034 | 0.002 |
| Green leafy vegetables | -0.027 | Nuts | 1 | -0.033 | 0.006 | Nuts | 1 | -0.033 | 0.001 |
| Artificial sweetened beverages | 0.008 | Fruits | 1 | -0.031 | 0.013 | Fruits | 1 | -0.033 | 0.000 |
| High-fat dairy | 0.008 | Beer | 1 | 0.028 | 0.010 | Tea | 1 | -0.024 | 0.001 |
| Other vegetables | -0.005 | Tea | 1 | -0.028 | 0.013 | Beer | 1 | 0.021 | 0.000 |
| Nuts | -0.004 | Oils | 0.8 | -0.013 | 0.011 | Oils | 1 | -0.008 | 0.000 |
| Wine | -0.004 | High-fat dairy | 0.8 | 0.007 | 0.014 | Other vegetables | 1 | -0.005 | 0.002 |
| Low-fat dairy | 0 | Artificial sweetened beverages | 0.8 | 0.006 | 0.010 | High-fat dairy | 1 | 0.003 | 0.000 |
| Butter | 0 | Poultry | 0.6 | 0.007 | 0.016 | Artificial sweetened beverages | 1 | 0.002 | 0.000 |

| | | | | | | | | | |
|---|---|---|---|---|---|---|---|---|---|
| Margarine | 0 | Other vegetables | 0.6 | -0.002 | 0.016 | Butter | 0 | 0 | 0 |
| Fruit juice | 0 | Butter | 0.3 | 0 | 0.001 | Coffee | 0 | 0 | 0 |
| Tomato | 0 | Coffee | 0.3 | 0 | 0.007 | Condiments | 0 | 0 | 0 |
| Cruciferous vegetables | 0 | Fish and other seafoods | 0.3 | 0 | 0.004 | Cruciferous vegetables | 0 | 0 | 0 |
| Dark yellow vegetables | 0 | Legumes | 0.3 | 0 | 0.000 | Dark yellow vegetables | 0 | 0 | 0 |
| Legumes | 0 | Mayonnaise | 0.3 | 0 | 0.003 | Eggs | 0 | 0 | 0 |
| Eggs | 0 | Garlic | 0.2 | 0 | 0 | Fish and other seafoods | 0 | 0 | 0 |
| Red meat | 0 | Cruciferous vegetables | 0.1 | 0 | 0 | Fries | 0 | 0 | 0 |
| Poultry | 0 | Fries | 0.1 | 0 | 0 | Fruit juice | 0 | 0 | 0 |
| Organ meat | 0 | Snack | 0.1 | 0 | 0 | Garlic | 0 | 0 | 0 |
| Fish and other seafoods | 0 | Soup | 0.1 | 0 | 0 | Legumes | 0 | 0 | 0 |
| Whole grains | 0 | Condiments | 0 | 0 | 0 | Low-fat dairy | 0 | 0 | 0 |
| Refined grains | 0 | Dark yellow vegetables | 0 | 0 | 0 | Margarine | 0 | 0 | 0 |
| Pizza | 0 | Eggs | 0 | 0 | 0 | Mayonnaise | 0 | 0 | 0 |
| Potato | 0 | Fruit juice | 0 | 0 | 0 | Pizza | 0 | 0 | 0 |
| Fries | 0 | Low-fat dairy | 0 | 0 | 0 | Potato | 0 | 0 | 0 |
| Snack | 0 | Margarine | 0 | 0 | 0 | Poultry | 0 | 0 | 0 |
| Beer | 0 | Pizza | 0 | 0 | 0 | Refined grains | 0 | 0 | 0 |
| Coffee | 0 | Potato | 0 | 0 | 0 | Red meat | 0 | 0 | 0 |
| Sweets | 0 | Refined grains | 0 | 0 | 0 | Snack | 0 | 0 | 0 |
| Soup | 0 | Red meat | 0 | 0 | 0 | Soup | 0 | 0 | 0 |

| | | | | | | | | | |
|---|---|---|---|---|---|---|---|---|---|
| Condiments | 0 | Sweets | 0 | 0 | 0 | Sweets | 0 | 0 | 0 |
| Garlic | 0 | Tomato | 0 | 0 | 0 | Tomato | 0 | 0 | 0 |
| Mayonnaise | 0 | Whole grains | 0 | 0 | 0 | Whole grains | 0 | 0 | 0 |
| Oils | 0 | Wine | 0 | 0 | 0 | Wine | 0 | 0 | 0 |

**Footnote:** The elastic-net model was used to identify food groups jointly associated with DunedinPoAm acceleration and to derive EDISEA component weights. Main model reports the fitted elastic-net coefficients from the primary specification. Cross-validation stability summarizes refits across cross-validation folds using the primary penalty selection rule. Selection frequency is the proportion of folds in which a food group's coefficient was non-zero. Estimate median and estimate IQR are the median and interquartile range of the non-zero coefficients across folds. Penalty parameter stability evaluates robustness to the choice of the regularization strength (λ) within the prespecified search grid. Selection frequency, median coefficient, and IQR are computed analogously across λ refits.

**Table S4. Correlations of EDISEA with multiple DNA methylation-based epigenetic aging biomarkers in HRS and NHANES**

| | HRS | | NHANES | |
|---|---|---|---|---|
| **Epigenetic clocks** | **N** | **ρ** | **N** | **ρ** |
| GrimAgeAccel | 1711 | **-0.323** | 2311 | **-0.222** |
| DunedinPoAm | 1711 | **-0.207** | 2311 | **-0.159** |
| Zhang AgeAccel | 1711 | **-0.187** | 2311 | -0.013 |
| PhenoAgeAccel | 1711 | **-0.125** | 2311 | **-0.085** |
| Hannum AgeAccel | 1711 | **-0.109** | 2311 | **-0.051** |
| Horvath AgeAccel | 1711 | **-0.104** | 2311 | **-0.061** |
| SkinBlood AgeAccel | 1711 | **-0.094** | 2311 | **-0.041** |
| Garagnani AgeAccel | 1711 | **-0.066** | - | - |
| Vidal-Bralo AgeAccel | 1711 | **-0.056** | 2311 | -0.013 |
| Weidner AgeAccel | 1711 | 0.018 | 2311 | 0.032 |
| Yang AgeAccel | 1711 | 0.017 | - | - |
| Lin AgeAccel | 1711 | -0.017 | 2311 | 0.015 |
| Bocklandt AgeAccel | 1711 | -0.010 | - | - |
| DNAmTL AgeAccel | - | - | 2311 | **0.100** |

**Footnote:** Spearman rank correlations (rho) between EDISEA and DNA methylation-based epigenetic aging biomarkers in HRS and NHANES. Measures are expressed as AgeAccel (age acceleration), defined as the residual from regressing the DNAm-derived age on chronological age. Higher values of GrimAgeAccel, DunedinPoAm, Zhang AgeAccel, PhenoAgeAccel, Hannum AgeAccel, Horvath AgeAccel, SkinBlood AgeAccel, Garagnani AgeAccel, Vidal-Bralo AgeAccel, Weidner AgeAccel, Yang AgeAccel, Lin AgeAccel, and Bocklandt AgeAccel indicate faster epigenetic aging. Higher values of DNAmTL AgeAccel indicate slower epigenetic aging.

**Table S5. Associations between EDISEA and all-cause mortality**

| | HRS | NHANES | UKB |
|---|---|---|---|
| Model 1 | **0.66 (0.60-0.74)** | **0.70 (0.66-0.74)** | **0.76 (0.74-0.79)** |
| Model 2 | **0.78 (0.69-0.89)** | **0.79 (0.74-0.84)** | **0.85 (0.82-0.88)** |
| Model 3 | **0.78 (0.69-0.88)** | **0.80 (0.75-0.85)** | **0.85 (0.82-0.89)** |

**Footnote:** HRs (95% CIs) for all-cause mortality estimated from Cox proportional hazards models, with EDISEA modeled continuously and scaled per $90^{th}$ to $10^{th}$ percentile difference of EDISEA. Model 1 adjusted for age and sex. Model 2 additionally adjusted for ethnicity, education, income, smoking, alcohol use, physical activity, body mass index, and total energy intake. Model 3 further adjusted for baseline comorbidities (diabetes, high blood pressure, and cardiovascular diseases).

**Table S6. Subgroup analyses of associations between EDISEA and all-cause mortality in HRS, NHANES and the UK Biobank**

| | HRS | | | NHANES | | | UKB | | |
|---|---|---|---|---|---|---|---|---|---|
| **Subgroup** | Cases/ Person -yrs | HR (95%) | P | Cases/ Person-yrs | HR (95%) | P | Cases/ Person-yrs | HR (95%) | P |
| **Age** | | | 0.039 | | | 0.009 | | | 0.189 |
| <65 | 206 / 30352 | **0.65 (0.53-0.80)** | | 1467 / 143546 | **0.78 (0.71-0.86)** | | 9245 / 2009246 | **0.84 (0.80-0.87)** | |
| >=65 | 1436 / 39026 | **0.85 (0.73-0.98)** | | 4235 / 87122 | 0.95 (0.85-1.06) | | 5807 / 377743 | **0.88 (0.83-0.93)** | |
| **Sex** | | | 0.015 | | | 0.839 | | | 0.699 |
| Male | 749 / 27833 | 0.88 (0.75-1.04) | | 3108 / 109671 | **0.85 (0.78-0.93)** | | 8817 / 1061233 | **0.85 (0.82-0.89)** | |
| Female | 893 / 41545 | **0.65 (0.54-0.79)** | | 2594 / 120997 | **0.84 (0.72-0.97)** | | 6235 / 1325756 | **0.84 (0.78-0.90)** | |
| **Education** | | | 0.498 | | | 0.751 | | | 0.071 |
| Below college | 958 / 33731 | **0.81 (0.69-0.95)** | | 4905 / 182485 | **0.85 (0.79-0.93)** | | 9619 / 1368129 | **0.87 (0.83-0.91)** | |
| College and above | 684 / 35647 | **0.75 (0.63-0.89)** | | 797 / 48182 | **0.83 (0.69-0.99)** | | 5433 / 1018860 | **0.81 (0.76-0.86)** | |
| **Income** | | | 0.880 | | | 0.965 | | | 0.0006 |
| Low | 670 / 18625 | **0.76 (0.64-0.91)** | | 1873 / 60384 | 0.86 (0.74-1.00) | | 7964 / 1308661 | **0.90 (0.86-0.95)** | |
| Medium | 528 / 17683 | **0.78 (0.62-0.97)** | | 2517 / 86948 | **0.85 (0.75-0.96)** | | 4845 / 767712 | **0.84 (0.79-0.89)** | |

| | | | | | | | | | |
|---|---|---|---|---|---|---|---|---|---|
| High | 444 / 33069 | 0.82 (0.65-1.02) | | 1312 / 83335 | **0.84 (0.74-0.95)** | | 2243 / 310615 | **0.75 (0.70-0.81)** | |
| **Smoking status** | | | 0.646 | | | 0.602 | | | 0.081 |
| Never | 674 / 31921 | **0.80 (0.65-0.99)** | | 2321 / 114189 | 0.87 (0.76-1.00) | | 6465 / 1370767 | **0.88 (0.82-0.93)** | |
| Former | 787 / 29981 | **0.80 (0.68-0.96)** | | 2323 / 76353 | 0.88 (0.76-1.01) | | 6671 / 838367 | **0.86 (0.82-0.90)** | |
| Current | 181 / 7476 | **0.70 (0.55-0.90)** | | 1058 / 40125 | **0.81 (0.72-0.91)** | | 1916 / 177855 | **0.78 (0.72-0.85)** | |
| **BMI, kg/m$^2$** | | | 0.028 | | | 0.734 | | | 0.019 |
| 0-24.9 | 601 / 18513 | **0.65 (0.55-0.78)** | | 1716 / 58379 | **0.84 (0.74-0.95)** | | 4552 / 914186 | **0.79 (0.74-0.85)** | |
| 25-29.9 | 558 / 25948 | 0.89 (0.72-1.10) | | 2079 / 85510 | **0.88 (0.78-0.99)** | | 6340 / 990620 | **0.85 (0.80-0.90)** | |
| >=30 | 483 / 24916 | 0.88 (0.70-1.11) | | 1907 / 86779 | **0.83 (0.73-0.93)** | | 4160 / 482183 | **0.90 (0.85-0.96)** | |

**Footnote:** Hazard ratios (HRs) and 95% confidence intervals (CIs) were estimated from Cox proportional hazards models for all-cause mortality within each subgroup, with EDISEA modeled continuously and scaled 90$^{th}$ to 10$^{th}$ percentile difference in EDISEA. Models were adjusted for the main analysis covariates in each cohort (including demographic factors, socioeconomic indicators, lifestyle factors, total energy intake). For NHANES, analyses accounted for the complex survey design. P values for interaction were obtained by adding a multiplicative EDISEA × subgroup term to the adjusted model and testing the interaction coefficient.

**Table S7. Sensitivity analyses of associations between EDISEA and all-cause mortality in HRS and the UK Biobank**

| | HRS | | | UK Biobank | | |
|---|---|---|---|---|---|---|
| **Analyses** | N | Cases | HR (95% CI) | N | Cases | HR (95% CI) |
| Primary analysis | 7398 | 1642 | **0.78 (0.69-0.89)** | 187035 | 15052 | **0.85 (0.82-0.88)** |
| Sensitivity 1 | 7204 | 1453 | **0.78 (0.69-0.89)** | 185731 | 13753 | **0.87 (0.83-0.90)** |
| Sensitivity 2 | 5851 | 1013 | **0.79 (0.67-0.92)** | 146869 | 12844 | **0.85 (0.81-0.88)** |

**Footnote:** HRs (95% CIs) for all-cause mortality were estimated using Cox proportional hazards models with EDISEA modeled continuously and scaled per the $90^{th}$ to $10^{th}$ percentile difference of EDISEA within each cohort. Sensitivity 1 excluded deaths occurring within the first 2 years of follow-up. Sensitivity 2 excluded participants with baseline functional limitation. In UK Biobank, baseline functional limitation was proxied using three EQ-5D-5L "health today" domains, mobility (Data-Field 120098), self-care (Data-Field 120099), and usual activities (Data-Field 120100), derived from the UK Biobank online follow-up questionnaire. Participants reporting any problem in any domain were classified as having limitation.

**Table S8. Plasma proteins with mediating effect between EDISEA and all-cause mortality**

| Plasma Proteins | Proportion | Pattern-Protein Beta | Protein-Mortality Beta |
|---|---|---|---|
| A1BG | 0.030* | -0.167*** | -0.059* |
| ACAA1 | 0.072*** | -0.269*** | -0.067*** |
| ACE2 | 0.228*** | -0.369*** | -0.136*** |
| ACP5 | 0.093*** | -0.353*** | -0.068*** |
| ACP6 | 0.026* | -0.147** | -0.052* |
| ACTN2 | -0.027*** | 0.143** | -0.066** |
| ADA2 | -0.079*** | 0.141** | -0.153*** |
| ADAM12 | 0.075*** | -0.167*** | -0.132*** |
| ADAMTS13 | 0.041* | 0.214*** | 0.048* |
| ADAMTS15 | 0.032* | -0.117* | -0.074*** |
| ADAMTS4 | 0.037* | -0.140** | -0.081*** |
| ADAMTS8 | 0.067*** | 0.273*** | 0.059** |
| ADAMTSL2 | 0.147*** | -0.277*** | -0.154*** |
| ADGRB3 | -0.041*** | 0.165*** | -0.075*** |
| ADGRD1 | 0.177*** | -0.491*** | -0.116*** |
| ADGRE2 | -0.036* | 0.139** | -0.063*** |
| ADGRE5 | -0.043*** | 0.143** | -0.079*** |
| ADGRF5 | 0.051*** | -0.227*** | -0.071** |
| ADGRG2 | 0.059*** | 0.160*** | 0.100*** |
| ADM | 0.316*** | -0.331*** | -0.248*** |
| AGRN | 0.071* | -0.108* | -0.156*** |
| AGXT | 0.049*** | -0.231*** | -0.057** |
| ALCAM | -0.139*** | 0.245*** | -0.146*** |
| ALPP | 0.061*** | -0.208*** | -0.077*** |
| AMBP | 0.083*** | -0.164*** | -0.129*** |
| ANG | 0.063*** | -0.195*** | -0.087*** |
| ANGPT2 | 0.075* | -0.111* | -0.179*** |
| ANGPTL2 | 0.054*** | -0.191*** | -0.069*** |
| ANGPTL4 | 0.067*** | -0.124** | -0.142*** |
| ANGPTL7 | 0.031* | -0.160*** | -0.049* |
| ANPEP | 0.057*** | -0.171*** | -0.085*** |
| APOM | -0.068* | -0.380*** | 0.047* |
| AREG | 0.106* | -0.123* | -0.223*** |
| ARSA | 0.031* | -0.175*** | -0.049* |
| ASGR1 | 0.233*** | -0.423*** | -0.157*** |
| ASGR2 | 0.084*** | -0.245*** | -0.113*** |
| B2M | 0.042*** | -0.136** | -0.098*** |
| BAIAP2 | 0.115*** | -0.262*** | -0.109*** |
| BCAM | -0.066*** | 0.170*** | -0.105*** |

| | | | |
|---|---|---|---|
| BCAN | 0.058*** | 0.218*** | 0.080*** |
| BCAT1 | 0.036* | -0.159** | -0.075*** |
| BMPER | 0.044* | -0.143** | -0.109*** |
| BOC | -0.064* | 0.269*** | -0.055** |
| BPIFB1 | 0.057*** | -0.161*** | -0.083*** |
| BST2 | 0.175*** | -0.320*** | -0.137*** |
| BTN2A1 | -0.056*** | 0.119* | -0.114*** |
| BTN3A2 | 0.046* | -0.116* | -0.107*** |
| C1QA | -0.041*** | 0.111* | -0.100*** |
| CA12 | -0.069*** | 0.200*** | -0.096*** |
| CA14 | 0.041* | 0.220*** | 0.050* |
| CA5A | 0.046*** | -0.164*** | -0.068*** |
| CA9 | -0.035* | 0.108* | -0.088*** |
| CALB1 | -0.052*** | 0.166*** | -0.086*** |
| CALB2 | 0.028* | -0.109* | -0.068*** |
| CALCA | 0.047*** | -0.104** | -0.134*** |
| CANT1 | 0.071*** | -0.224*** | -0.084*** |
| CCDC80 | 0.057*** | -0.139*** | -0.110*** |
| CCL14 | 0.066*** | -0.133** | -0.123*** |
| CCL16 | 0.119*** | -0.258*** | -0.117*** |
| CCL17 | 0.081*** | -0.179*** | -0.110*** |
| CCL20 | 0.098*** | -0.228*** | -0.112*** |
| CCL21 | 0.040* | -0.121* | -0.087*** |
| CCL22 | 0.083*** | -0.220*** | -0.098*** |
| CCL23 | 0.063*** | -0.176*** | -0.092*** |
| CCL27 | 0.027* | -0.142** | -0.051* |
| CCL28 | 0.031* | -0.164*** | -0.051** |
| CCL3 | 0.072*** | -0.193*** | -0.101*** |
| CCL5 | 0.055*** | -0.192*** | -0.074*** |
| CCL7 | 0.052* | -0.142** | -0.104*** |
| CCL8 | 0.037* | -0.176*** | -0.056** |
| CCN2 | 0.031* | -0.175*** | -0.047* |
| CCN3 | -0.053* | 0.104* | -0.133*** |
| CD14 | 0.215*** | -0.384*** | -0.131*** |
| CD274 | 0.075*** | -0.161*** | -0.138*** |
| CD276 | -0.063*** | 0.127** | -0.130*** |
| CD300A | 0.056*** | -0.174*** | -0.113*** |
| CD302 | 0.079*** | -0.127** | -0.157*** |
| CD40 | 0.027* | -0.114* | -0.061*** |
| CD93 | -0.095*** | 0.290*** | -0.089*** |
| CD99L2 | 0.157*** | -0.445*** | -0.097*** |
| CDCP1 | 0.083*** | -0.165*** | -0.145*** |

| | | | |
|---|---|---|---|
| CDH2 | 0.071*** | -0.178*** | -0.111*** |
| CDH6 | -0.062*** | 0.231*** | -0.071*** |
| CDHR2 | 0.088* | -0.328*** | -0.074** |
| CEACAM1 | 0.123*** | -0.317*** | -0.088*** |
| CEACAM16 | 0.118* | 0.580*** | 0.063** |
| CEACAM5 | 0.084*** | -0.130** | -0.138*** |
| CGREF1 | 0.084*** | -0.269*** | -0.082*** |
| CHAD | 0.026* | 0.141** | 0.061** |
| CHCHD10 | 0.071* | -0.118* | -0.193*** |
| CHCHD6 | 0.073*** | -0.240*** | -0.098*** |
| CHI3L1 | 0.434*** | -0.633*** | -0.145*** |
| CKB | -0.093*** | 0.209*** | -0.143*** |
| CLEC3B | 0.055* | 0.132* | 0.145*** |
| CLEC4D | 0.087*** | -0.174*** | -0.134*** |
| CLEC5A | 0.057*** | -0.139** | -0.113*** |
| CLEC6A | 0.098*** | -0.197*** | -0.121*** |
| CNDP1 | -0.066*** | -0.253*** | 0.069*** |
| CNTN5 | 0.064*** | 0.281*** | 0.064** |
| COCH | 0.069*** | -0.325*** | -0.067** |
| COL18A1 | 0.150*** | -0.185*** | -0.193*** |
| COL4A1 | -0.070*** | 0.130** | -0.141*** |
| COQ7 | 0.041*** | -0.233*** | -0.057** |
| CPM | 0.090*** | -0.238*** | -0.108*** |
| CPXM1 | 0.047*** | -0.188*** | -0.058** |
| CR1 | -0.021* | 0.134** | -0.056* |
| CRELD1 | 0.079*** | -0.230*** | -0.118*** |
| CRHBP | 0.040*** | -0.151*** | -0.069*** |
| CRTAC1 | 0.040*** | 0.165*** | 0.062** |
| CSPG4 | -0.030*** | 0.157*** | -0.066** |
| CST7 | 0.027* | -0.120* | -0.057*** |
| CSTB | 0.079*** | -0.146** | -0.128*** |
| CTSC | 0.067*** | -0.210*** | -0.078*** |
| CTSD | 0.195*** | -0.483*** | -0.089*** |
| CTSL | -0.098*** | 0.202*** | -0.133*** |
| CXADR | 0.112*** | -0.229*** | -0.127*** |
| CXCL10 | 0.070*** | -0.176*** | -0.102*** |
| CXCL11 | 0.074*** | -0.183*** | -0.107*** |
| CXCL16 | 0.060* | -0.125** | -0.121*** |
| CXCL17 | 0.099*** | -0.143*** | -0.173*** |
| CXCL8 | 0.043*** | -0.138** | -0.090*** |
| CXCL9 | 0.091*** | -0.150*** | -0.146*** |
| DBH | 0.022*** | 0.147** | 0.053** |

| | | | |
|---|---|---|---|
| DCBLD2 | -0.040* | 0.110* | -0.084*** |
| DCTPP1 | -0.029* | 0.121* | -0.066*** |
| DKK1 | 0.020*** | -0.111* | -0.053** |
| DLL1 | -0.084*** | 0.143** | -0.141*** |
| DPY30 | 0.032* | -0.098* | -0.093*** |
| DRAXIN | -0.024* | 0.104* | -0.067*** |
| DSC2 | -0.121*** | 0.245*** | -0.137*** |
| EBI3_IL27 | -0.063* | 0.114* | -0.146*** |
| ECHDC3 | 0.085*** | -0.322*** | -0.080*** |
| EFNA1 | 0.062*** | -0.108* | -0.158*** |
| EGFL7 | 0.044* | -0.114* | -0.096*** |
| EGLN1 | 0.018* | -0.118* | -0.042* |
| ENPP2 | -0.031* | 0.088* | -0.089*** |
| ENPP6 | 0.044* | 0.250*** | 0.059* |
| ENPP7 | 0.092*** | -0.330*** | -0.073*** |
| EPHA1 | 0.026* | -0.105* | -0.063** |
| EPO | -0.061*** | 0.181*** | -0.092*** |
| EPS8L2 | 0.040* | -0.114* | -0.093*** |
| F10 | -0.052* | -0.341*** | 0.051* |
| F3 | -0.026* | 0.116* | -0.064** |
| F9 | 0.108*** | -0.444*** | -0.064** |
| FABP1 | 0.077*** | -0.130** | -0.143*** |
| FABP4 | 0.199*** | -0.297*** | -0.164*** |
| FAM20A | 0.098*** | -0.412*** | -0.074*** |
| FAM3C | 0.062* | -0.117* | -0.139*** |
| FCAMR | -0.058*** | 0.194*** | -0.099*** |
| FCAR | 0.030*** | -0.154*** | -0.053** |
| FCER2 | -0.027* | 0.129** | -0.058*** |
| FCN1 | 0.044*** | -0.199*** | -0.076** |
| FCN2 | -0.016* | -0.101* | 0.043* |
| FGF21 | 0.251*** | -0.697*** | -0.091*** |
| FGF5 | -0.033* | 0.112* | -0.077*** |
| FGFBP1 | 0.071*** | 0.192*** | 0.085*** |
| FGFR2 | 0.075*** | -0.143*** | -0.118*** |
| FGL1 | 0.055*** | -0.125** | -0.144*** |
| FLRT2 | -0.062*** | 0.227*** | -0.080*** |
| FLT1 | -0.022* | 0.119* | -0.049* |
| FOLR1 | -0.130*** | 0.255*** | -0.118*** |
| FST | 0.071* | -0.339*** | -0.055** |
| FSTL3 | 0.253*** | -0.316*** | -0.194*** |
| FUOM | 0.028* | -0.174*** | -0.054* |
| FUT3_FUT5 | 0.066*** | -0.180*** | -0.094*** |

| | | | |
|---|---|---|---|
| GALNT10 | 0.094*** | -0.202*** | -0.126*** |
| GDF15 | 0.166*** | -0.147*** | -0.281*** |
| GGT1 | 0.244*** | -0.514*** | -0.127*** |
| GIPC2 | 0.043* | -0.115* | -0.114*** |
| GLA | 0.069*** | -0.289*** | -0.081*** |
| GOLM2 | 0.094*** | -0.243*** | -0.097*** |
| GPR37 | 0.131*** | -0.235*** | -0.128*** |
| GPRC5C | 0.055*** | -0.165*** | -0.104*** |
| GRN | 0.056*** | -0.169*** | -0.091*** |
| GRPEL1 | 0.133*** | -0.321*** | -0.113*** |
| GSN | 0.028*** | 0.152** | 0.056*** |
| GUCA2A | -0.140*** | 0.330*** | -0.111*** |
| HAO1 | 0.059* | -0.347*** | -0.047* |
| HAVCR1 | 0.235*** | -0.296*** | -0.205*** |
| HEG1 | 0.031* | -0.132** | -0.082*** |
| HGF | 0.214*** | -0.292*** | -0.186*** |
| HMOX2 | 0.067*** | -0.200*** | -0.086*** |
| HNMT | 0.039*** | -0.154*** | -0.074*** |
| HPSE | 0.035* | -0.190*** | -0.055* |
| HRC | -0.034* | 0.143** | -0.081*** |
| HS3ST3B1 | 0.027* | -0.122* | -0.058** |
| HS6ST2 | -0.050*** | 0.203*** | -0.073*** |
| HYAL1 | 0.117*** | -0.290*** | -0.100*** |
| HYOU1 | 0.058*** | -0.179*** | -0.082*** |
| ICAM1 | 0.105*** | -0.187*** | -0.152*** |
| IDUA | 0.030*** | -0.146** | -0.056** |
| IFNLR1 | 0.045*** | -0.190*** | -0.065*** |
| IGDCC4 | 0.062*** | 0.312*** | 0.064** |
| IGF2R | 0.067*** | -0.233*** | -0.078*** |
| IGFBP2 | -0.131*** | 0.212*** | -0.147*** |
| IGFBP4 | 0.209*** | -0.253*** | -0.198*** |
| IGFBPL1 | 0.148*** | -0.252*** | -0.150*** |
| IGSF21 | 0.028* | 0.120* | 0.077*** |
| IGSF3 | 0.047*** | -0.116* | -0.109*** |
| IL10RB | 0.076*** | -0.147*** | -0.134*** |
| IL15 | 0.131*** | -0.270*** | -0.130*** |
| IL17C | 0.054* | -0.146** | -0.093*** |
| IL18 | 0.058*** | -0.206*** | -0.070*** |
| IL18R1 | 0.076*** | -0.169*** | -0.116*** |
| IL19 | 0.063*** | -0.129** | -0.125*** |
| IL1R1 | -0.070*** | 0.158*** | -0.128*** |
| IL1RL1 | 0.093*** | -0.183*** | -0.129*** |

| | | | |
|---|---|---|---|
| IL1RN | 0.082*** | -0.216*** | -0.094*** |
| IL22 | 0.037* | -0.135** | -0.088*** |
| IL6 | 0.105*** | -0.215*** | -0.133*** |
| IMMT | 0.090*** | -0.256*** | -0.113*** |
| INHBB | 0.060*** | -0.132** | -0.152*** |
| INSL3 | 0.020* | 0.066* | 0.094* |
| INSL5 | 0.061*** | -0.261*** | -0.072*** |
| ITGA11 | 0.078*** | 0.194*** | 0.111*** |
| ITGA5 | 0.060*** | -0.138** | -0.117*** |
| ITGAM | 0.048*** | 0.189*** | 0.070*** |
| ITGBL1 | 0.052* | -0.140** | -0.117*** |
| JAM2 | -0.070*** | 0.189*** | -0.103*** |
| KDR | -0.039* | -0.186*** | 0.052** |
| KIT | 0.049*** | 0.180*** | 0.066*** |
| KITLG | 0.057* | 0.298*** | 0.051** |
| KLK10 | 0.028* | -0.116* | -0.056*** |
| KLKB1 | -0.041* | -0.156** | 0.085*** |
| KLRD1 | 0.021* | -0.103* | -0.053** |
| KRT18 | 0.038* | -0.115** | -0.091*** |
| LAMP3 | 0.084*** | -0.124** | -0.163*** |
| LBP | 0.035* | -0.132** | -0.072*** |
| LBR | 0.026* | -0.160*** | -0.044* |
| LETM1 | 0.024* | -0.125* | -0.066*** |
| LGALS1 | 0.061*** | -0.147*** | -0.114*** |
| LGALS3BP | 0.062*** | -0.212*** | -0.095*** |
| LGALS9 | 0.099*** | -0.137** | -0.195*** |
| LGMN | 0.043*** | -0.148** | -0.077*** |
| LILRA5 | 0.053*** | -0.152*** | -0.097*** |
| LILRB4 | 0.151*** | -0.242*** | -0.157*** |
| LMNB2 | 0.067*** | -0.124* | -0.173*** |
| LRRN1 | 0.101*** | 0.222*** | 0.112*** |
| LTBP3 | 0.037* | -0.122* | -0.075*** |
| LY6D | 0.048* | -0.126** | -0.093*** |
| LY96 | 0.045*** | -0.157*** | -0.087*** |
| MAD1L1 | 0.044*** | -0.149*** | -0.093*** |
| MANSC1 | -0.038*** | 0.162*** | -0.060** |
| MARCO | 0.046*** | -0.180*** | -0.067*** |
| MB | -0.028* | 0.114** | -0.064** |
| MDK | 0.122*** | -0.256*** | -0.111*** |
| MME | 0.154*** | -0.410*** | -0.101*** |
| MMP1 | 0.056*** | -0.229*** | -0.058** |
| MMP10 | 0.048*** | -0.191*** | -0.070*** |

| | | | |
|---|---|---|---|
| MMP7 | 0.162*** | -0.188*** | -0.206*** |
| MMP8 | 0.056*** | -0.223*** | -0.068*** |
| MMP9 | 0.068*** | -0.258*** | -0.074*** |
| MRC1 | 0.093*** | -0.224*** | -0.138*** |
| MSR1 | 0.109*** | -0.175*** | -0.160*** |
| MSTN | 0.045* | 0.196*** | 0.055* |
| MUC13 | 0.069*** | -0.191*** | -0.091*** |
| MVK | 0.067*** | -0.252*** | -0.068*** |
| MYBPC1 | -0.035*** | 0.120* | -0.097*** |
| MYL3 | -0.029* | 0.130** | -0.076*** |
| MZB1 | 0.126*** | -0.255*** | -0.118*** |
| NCR1 | -0.046*** | 0.171*** | -0.075*** |
| NELL1 | 0.113*** | 0.305*** | 0.086*** |
| NHLRC3 | 0.116*** | -0.370*** | -0.104*** |
| NID1 | 0.084*** | -0.267*** | -0.084*** |
| NME3 | 0.032* | -0.199*** | -0.042* |
| NOMO1 | 0.069* | -0.322*** | -0.061** |
| NOTCH1 | -0.067* | 0.328*** | -0.052** |
| NOTCH3 | -0.080*** | 0.188*** | -0.101*** |
| NPL | 0.044*** | -0.173*** | -0.083*** |
| NRCAM | -0.076*** | 0.239*** | -0.088*** |
| NRP2 | -0.035* | 0.104* | -0.094*** |
| NT5C1A | 0.057* | -0.125** | -0.143*** |
| NT5E | 0.067*** | -0.222*** | -0.073*** |
| OCLN | 0.099*** | -0.146** | -0.216*** |
| OMG | 0.036*** | 0.147** | 0.065*** |
| ORM1 | 0.069*** | -0.319*** | -0.072*** |
| OSM | 0.077*** | -0.269*** | -0.078*** |
| PALM2 | 0.178*** | -0.261*** | -0.212*** |
| PCBD1 | 0.032* | -0.199*** | -0.056* |
| PCDH17 | -0.050*** | 0.189*** | -0.072*** |
| PCSK9 | 0.118*** | -0.345*** | -0.087*** |
| PDCD1 | -0.058* | 0.120* | -0.131*** |
| PDCD1LG2 | -0.029* | 0.113* | -0.072*** |
| PDZK1 | 0.063*** | -0.191*** | -0.100*** |
| PEPD | -0.019* | -0.119* | 0.056** |
| PGLYRP1 | 0.055*** | -0.134** | -0.100*** |
| PHLDB1 | 0.025*** | -0.127* | -0.066** |
| PI3 | 0.066*** | -0.132** | -0.133*** |
| PIGR | 0.197*** | -0.246*** | -0.199*** |
| PILRA | 0.056* | -0.106* | -0.141*** |
| PKD1 | -0.034* | 0.121* | -0.092*** |

| | | | |
|---|---|---|---|
| PLA2G15 | 0.142*** | -0.274*** | -0.114*** |
| PLAT | 0.120*** | -0.367*** | -0.079*** |
| PLAU | -0.036*** | 0.150** | -0.068*** |
| PLAUR | 0.149*** | -0.169*** | -0.212*** |
| PLXDC1 | 0.034* | 0.121* | 0.068** |
| POLR2F | 0.037*** | -0.131** | -0.078*** |
| PPL | 0.025* | -0.132* | -0.060** |
| PRAP1 | 0.135*** | -0.315*** | -0.139*** |
| PRCP | 0.054*** | -0.195*** | -0.070*** |
| PRND | -0.070*** | 0.305*** | -0.073*** |
| PRSS2 | 0.074*** | -0.345*** | -0.052** |
| PRSS8 | 0.497*** | -0.498*** | -0.221*** |
| PTGDS | -0.050* | 0.107* | -0.136*** |
| PTN | 0.078*** | -0.126** | -0.147*** |
| PTX3 | 0.050*** | -0.138** | -0.093*** |
| PVR | 0.143*** | -0.377*** | -0.098*** |
| RARRES2 | 0.113*** | -0.288*** | -0.106*** |
| RBP5 | 0.117*** | -0.272*** | -0.115*** |
| RBP7 | 0.022* | -0.134** | -0.054** |
| REG4 | 0.124*** | -0.206*** | -0.156*** |
| REN | 0.141*** | -0.222*** | -0.160*** |
| RGMB | -0.033* | 0.200*** | -0.045* |
| RNASE1 | 0.078*** | -0.181*** | -0.143*** |
| RNASE4 | 0.070*** | -0.199*** | -0.127*** |
| RNASE6 | 0.082*** | -0.198*** | -0.126*** |
| ROBO2 | -0.053*** | 0.167*** | -0.088*** |
| ROR1 | -0.050*** | 0.218*** | -0.064*** |
| RSPO1 | 0.047*** | -0.125** | -0.111*** |
| SCARB2 | 0.087*** | -0.130** | -0.179*** |
| SCARF1 | 0.046*** | -0.130** | -0.086*** |
| SCARF2 | -0.111*** | 0.177*** | -0.159*** |
| SCPEP1 | 0.024* | -0.130* | -0.062** |
| SDHB | 0.031* | -0.204*** | -0.050* |
| SELE | 0.061* | -0.281*** | -0.056** |
| SELENOP | 0.024* | 0.114* | 0.068** |
| SEPTIN8 | 0.063*** | -0.158*** | -0.131*** |
| SERPINA5 | -0.034*** | -0.186*** | 0.064** |
| SERPINA9 | 0.044*** | -0.214*** | -0.055** |
| SERPINF2 | 0.022* | 0.113* | 0.061** |
| SEZ6L2 | -0.060*** | 0.141*** | -0.115*** |
| SIAE | 0.040*** | -0.157*** | -0.061** |
| SIGLEC7 | 0.073*** | -0.225*** | -0.088*** |

| | | | |
|---|---|---|---|
| SLAMF7 | 0.049*** | -0.139** | -0.090*** |
| SLAMF8 | 0.094*** | -0.181*** | -0.124*** |
| SLC39A5 | 0.091*** | -0.291*** | -0.083*** |
| SMPD1 | 0.109*** | -0.325*** | -0.087*** |
| SNCG | 0.052*** | -0.187*** | -0.086*** |
| SORT1 | 0.060*** | -0.286*** | -0.057** |
| SPON2 | 0.103*** | -0.158*** | -0.179*** |
| SSC4D | 0.037*** | 0.118** | 0.079*** |
| ST6GAL1 | 0.120*** | -0.224*** | -0.137*** |
| STAB2 | 0.031* | -0.178*** | -0.056* |
| STC1 | 0.176*** | -0.441*** | -0.100*** |
| SULT2A1 | 0.062*** | -0.249*** | -0.066*** |
| SUSD4 | -0.043*** | 0.167*** | -0.083*** |
| TACSTD2 | 0.036* | -0.122* | -0.073*** |
| TCTN3 | 0.065* | -0.326*** | -0.063** |
| TFF3 | 0.076* | -0.117* | -0.171*** |
| TFRC | -0.050*** | 0.153*** | -0.087*** |
| TGFA | 0.124*** | -0.224*** | -0.148*** |
| TGFB1 | 0.081*** | -0.253*** | -0.084*** |
| THBD | -0.029*** | 0.131** | -0.063*** |
| THBS2 | 0.131*** | -0.243*** | -0.139*** |
| THBS4 | -0.022* | 0.105* | -0.056** |
| TIMP1 | 0.133*** | -0.194*** | -0.186*** |
| TIMP4 | 0.155*** | -0.354*** | -0.114*** |
| TMPRSS11D | 0.027* | -0.108* | -0.077*** |
| TNFRSF10A | 0.176*** | -0.294*** | -0.159*** |
| TNFRSF10B | 0.036* | -0.099* | -0.107*** |
| TNFRSF11B | 0.148*** | -0.240*** | -0.149*** |
| TNFRSF12A | 0.119*** | -0.148*** | -0.183*** |
| TNFRSF19 | -0.064*** | 0.131** | -0.133*** |
| TNFRSF6B | 0.214*** | -0.386*** | -0.154*** |
| TNFSF11 | 0.057*** | 0.228*** | 0.066*** |
| TNFSF13B | 0.101*** | -0.168*** | -0.159*** |
| TNR | 0.044*** | 0.184*** | 0.069*** |
| TPP1 | 0.206*** | -0.436*** | -0.114*** |
| TREM2 | 0.126*** | -0.253*** | -0.124*** |
| TSPAN1 | -0.024*** | 0.113* | -0.050*** |
| TTR | -0.056*** | -0.189*** | 0.105*** |
| TXNDC15 | 0.057*** | -0.193*** | -0.070*** |
| UMOD | 0.112*** | 0.231*** | 0.118*** |
| VAMP5 | 0.058*** | -0.190*** | -0.076*** |
| VCAM1 | -0.062* | 0.113* | -0.140*** |

| | | | |
|---|---|---|---|
| VEGFA | 0.125*** | -0.260*** | -0.120*** |
| VEGFB | 0.026* | -0.134** | -0.059** |
| VMO1 | 0.029* | -0.131** | -0.061** |
| VSIG4 | 0.101*** | -0.161*** | -0.170*** |
| VSNL1 | 0.041*** | -0.174*** | -0.074*** |
| VWA1 | 0.183*** | -0.512*** | -0.095*** |
| WFIKKN1 | 0.051*** | 0.186*** | 0.082*** |
| XPNPEP2 | -0.039* | -0.203*** | 0.047* |
| YAP1 | 0.071* | -0.117** | -0.191*** |

**Table S9. Associations between EDISEA and brain cortical thickness**

| | Left | | | Right | | |
|---|---|---|---|---|---|---|
| **Regions** | Beta | P value | FDR | Beta | P value | FDR |
| GlobalMeanMean thickness | 0.034 | **0.003** | **0.018** | 0.023 | **0.040** | 0.079 |
| bankssts | 0.037 | **0.002** | **0.016** | 0.030 | **0.013** | **0.038** |
| caudalanteriorcingulate | 0.006 | 0.639 | 0.725 | 0.012 | 0.327 | 0.412 |
| caudalmiddlefrontal | 0.028 | **0.015** | **0.040** | 0.028 | **0.014** | **0.038** |
| cuneus | 0.006 | 0.622 | 0.716 | -0.002 | 0.887 | 0.914 |
| entorhinal | 0.042 | **0.000** | **0.008** | 0.017 | 0.137 | 0.222 |
| frontalpole | 0.013 | 0.264 | 0.345 | 0.017 | 0.163 | 0.251 |
| fusiform | 0.030 | **0.012** | **0.038** | 0.019 | 0.114 | 0.189 |
| inferiorparietal | 0.027 | **0.016** | **0.040** | 0.030 | **0.008** | **0.031** |
| inferiortemporal | 0.016 | 0.166 | 0.251 | 0.014 | 0.238 | 0.318 |
| insula | 0.039 | **0.001** | **0.012** | 0.033 | **0.006** | **0.029** |
| isthmuscingulate | 0.030 | **0.012** | **0.038** | 0.032 | **0.006** | **0.029** |
| lateraloccipital | -0.001 | 0.938 | 0.938 | -0.011 | 0.339 | 0.420 |
| lateralorbitofrontal | 0.022 | 0.076 | 0.136 | 0.010 | 0.421 | 0.511 |
| lingual | 0.005 | 0.667 | 0.738 | 0.015 | 0.223 | 0.309 |
| medialorbitofrontal | 0.029 | **0.017** | **0.040** | 0.015 | 0.212 | 0.300 |
| middletemporal | 0.039 | **0.001** | **0.012** | 0.027 | **0.024** | 0.055 |
| paracentral | 0.032 | **0.007** | **0.030** | 0.036 | **0.002** | **0.016** |
| parahippocampal | 0.030 | **0.014** | **0.038** | 0.020 | 0.088 | 0.149 |
| parsopercularis | 0.024 | **0.038** | 0.078 | 0.026 | **0.026** | 0.060 |
| parsorbitalis | 0.017 | 0.148 | 0.234 | 0.005 | 0.673 | 0.738 |
| parstriangularis | 0.002 | 0.836 | 0.888 | -0.003 | 0.803 | 0.867 |
| pericalcarine | -0.001 | 0.936 | 0.938 | -0.017 | 0.175 | 0.259 |
| postcentral | 0.012 | 0.293 | 0.376 | 0.015 | 0.207 | 0.300 |
| posteriorcingulate | 0.039 | **0.001** | **0.012** | 0.033 | **0.006** | **0.029** |
| precentral | 0.030 | **0.011** | **0.038** | 0.034 | **0.003** | **0.018** |
| precuneus | 0.023 | **0.041** | 0.079 | 0.007 | 0.548 | 0.654 |
| rostralanteriorcingulate | 0.031 | **0.009** | **0.034** | 0.002 | 0.852 | 0.891 |
| rostralmiddlefrontal | 0.021 | 0.064 | 0.118 | 0.006 | 0.614 | 0.716 |
| superiorfrontal | 0.049 | **0.000** | **0.001** | 0.041 | **0.000** | **0.005** |
| superiorparietal | 0.020 | 0.078 | 0.136 | 0.014 | 0.233 | 0.317 |
| superiortemporal | 0.043 | **0.000** | **0.005** | 0.028 | **0.013** | **0.038** |
| supramarginal | 0.038 | **0.001** | **0.010** | 0.022 | 0.055 | 0.105 |
| transversetemporal | 0.026 | **0.034** | 0.072 | 0.026 | **0.033** | 0.071 |

**Table S10. ICD-10 defined outcomes in the UK Biobank**

| Category | Disease | ICD-10 |
|---|---|---|
| Blood, blood-forming organs and certain immune disorders | Anemia | D50-D64 |
| | Blood, blood-forming organs and certain immune disorders | D50-D89 |
| Cancer | Cancer | C00-C97 |
| | Breast cancer | C50 |
| | Colorectal cancer | C18-C20 |
| | Leukemia | C81-C96 |
| | Lung cancer | C34 |
| | Melanoma | C43-C44 |
| | Oesophagus cancer | C15 |
| | Prostate cancer | C61 |
| | Stomach cancer | C16 |
| Circulatory system disorders | Circulatory system disorders | I00-I99 |
| | Arrhythmias | I46-I49 |
| | Cerebrovascular diseases | I60-I69 |
| | Heart failure | I50 |
| | Hypertension | I10-I15 |
| | Ischaemic heart diseases | I20-I25 |
| | Peripheral artery diseases | I70-I79 |
| | Stroke | I60-I61; I63-I64 |
| Digestive system disorders | Digestive system disorders | K00-K93 |
| | Inflammatory bowel disease | K50-K52 |
| | Liver diseases | K70-K77 |
| | Pancreatitis | K85 |
| Ear and mastoid process disorders | Ear and mastoid process disorders | H60-H99 |
| Endocrine, nutritional and metabolic diseases | Endocrine, nutritional and metabolic diseases | E00-E90 |
| | Diabetes | E10-E14 |
| | Obesity | E66 |
| Eye and adnexa disorders | Eye and adnexa disorders | H00-H59 |
| | Cataracts | H25-H26 |
| | Glaucoma | H40 |
| Genitourinary system disorders | Genitourinary system disorders | N00-N99 |
| | Renal failure | N17-N19 |
| Certain infectious and parasitic diseases | Certain infectious and parasitic diseases | A01-B89 |
| | Bacterial infections | A01-A79 |
| | Viral infections | A80-B34 |

| | | |
|---|---|---|
| Musculoskeletal system and connective tissue disorders | Musculoskeletal system and connective tissue disorders | M00-M99 |
| | Osteoarthritis | M15-M19 |
| | Osteoporosis | M80-M82 |
| | Rheumatoid arthritis disorders | M05-M06; M08; M13; M30-M35; M45 |
| | Sciatica | M50-M51 |
| | Soft tissue disorders | M60-M79 |
| Mental and behavioural disorders | Mental and behavioural disorders | F00-F99 |
| | Dementia | F00-F03; G30; G31 |
| | Mood disorders | F30-F39 |
| | Neurotic disorders | F40-F48 |
| | Psychotic disorders | F20-F29 |
| | Substance abuse | F10-F19 |
| Nervous system disorders | Nervous system disorders | G00-G99 |
| | Epilepsy | G40-G42 |
| | Parkinson's disease | G20-G22 |
| | Sleep disorders | G47 |
| | Transient ischaemic attack | G45-G46 |
| Respiratory system disorders | Respiratory system disorders | J00-J99 |
| | Asthma | J45-J46 |
| | Chronic obstructive pulmonary disease | J40-J44; J47 |
| | Influenza and Pneumonia | J09-J18 |
| Skin and subcutaneous tissue disorders | Skin and subcutaneous tissue disorders | L00-L99 |
| | Infections and eczema | L00-L08; L20-L30 |

**Table S11. Missing rates of covariates**

| | Missing Rates, % | | | |
|---|---|---|---|---|
| **Characteristics** | **HRS** | **NHANES** | **ELSA** | **UKB** |
| Age | 0 | 0 | 0 | 0 |
| Sex | 0 | 0 | 0 | 0 |
| Ethnicity | 0 | 0 | 0 | 0.36 |
| Education | 0 | 0.18 | 7.75 | 0.49 |
| Income | 5.56 | 8.88 | 0.90 | 0.13 |
| Smoking status | 0 | 0.09 | 0.02 | 0.26 |
| Alcohol consumption | 5.99 | 37.67 | 2.45 | 0.10 |
| Physical activity | 0 | 20.16 | 0 | 17.39 |
| Body mass index | 0 | 1.89 | 16.55 | 0.28 |
| Total energy intake | 0 | 0 | 0 | 0 |
| Self-reported health conditions | | | | |
| Diabetes | 0 | 0 | 0 | 0.21 |
| High blood pressure | 0 | 10.71 | 0 | 0.16 |
| Cardiovascular diseases | 0 | 0.01 | 0 | 0.16 |
